# Modeling the effectiveness of Esperanza Window Traps as a complementary vector control strategy for achieving the community-wide elimination of Onchocerciasis

**DOI:** 10.1101/2024.10.25.24316075

**Authors:** Shakir Bilal, Morgan E. Smith, Swarnali Sharma, Wajdi Zaatour, Ken Newcomb, Thomas R. Unnasch, Edwin Michael

## Abstract

Mathematical models of parasite transmission provide powerful quantitative tools for evaluating the impact of interventions for bringing about the control or elimination of community-level disease transmission. A key attribute of such tools is that they allow integration of field observations regarding the effectiveness of an intervention with the processes of parasite transmission in communities to allow the exploration of parameters connected with the optimal deployment of the intervention to meet various community-wide control or elimination goals. In this work, we analyze the effectiveness of the Esperanza Window Trap (EWT), a recently developed black fly control tool, for eliminating the transmission of *Onchocera volvulus* in endemic settings by coupling seasonally-driven onchocerciasis transmission models identified for representative villages in Uganda with a landscape-level, spatially-informed model of EWT trap configurations for reducing Simulid fly populations in a given endemic setting. Our results indicate that when EWT traps are used in conjunction with MDA programs there are significant savings in the number of years needed to reach a specified set of elimination targets compared to the use of MDA alone. Adding EWT after the meeting of these thresholds and stoppage of MDA also significantly enhances the long-term sustained elimination of onchocerciasis. The number of traps required is driven by the trap black fly killing efficiency, capture range, desired coverage, inter-trap distance, size of location, and the spatial heterogeneity obtaining for the fly population in a given village/site. These findings provide important new knowledge regarding the feasibility and effectiveness of the community-wide use of EWT as a supplementary intervention alongside MDA for accelerating and sustaining the achievement of sustainable onchocerciasis elimination. Our coupling of landscape models of EWT deployment with the seasonal onchocerciasis transmission model also highlights how population-level macroparasite models may be extended effectively for modeling the effects of spatio-temporal processes on control efforts.

**Author summary:** While empirical studies have highlighted the effectiveness of the Esperanza Window Trap (EWT) as a potential tool for reducing biting black fly populations, information regarding how to implement these traps in the field to bring about community-wide elimination of onchocerciasis transmission is lacking. Here, we show how coupling a data-driven seasonal onchocerciasis transmission model with a landscape model of EWT trap networks can provide a flexible and powerful quantitative framework for addressing the effectiveness of deploying EWT in the field for bringing about parasite elimination in conjunction with mass drug administration (MDA). Our results demonstrate that including EWT traps with ivermectin MDA can significantly reduce timelines to reach elimination thresholds, while introducing these traps post-MDA can ensure the sustained long-term elimination of parasite transmission. The optimal trap configuration for meeting these goals will depend on the trap efficiencies for fly capture and killing, trap attractant range, field coverage, inter-trap distance, number of traps used, area of a control setting and the spatial variation observed for the density of biting black flies. This work also highlights how population-level models of macroparasite transmission dynamics could be extended successfully to effectively investigate these questions.

## Introduction

Onchocerciasis (river blindness), caused by infection with the *Simulium* black fly-transmitted filarial nematode, *Onchocerca volvulus,* continues to remain a serious tropical disease in many parts of Africa and in certain settings in Latin America (1–3). Current estimates, for example, suggest that at least 14.65 million people may be infected with the parasite, and that up to 217.5 million people continue to be at risk of infection in these regions (1, 4–7). This is despite the institution of long-term large regional and national-scale control programs, beginning with the initiation of vector control-based efforts from the mid-1970s in West Africa before community-wide mass drug administration (MDA) of ivermectin became the mainstay of national interventions from the 1990s (5, 8–10). While the initial focus of these programs was on morbidity control (11–13), the success of using MDA alone in achieving the elimination of onchocerciasis as a public health problem in 11 out of 13 foci in the Americas (13, 14), and in interrupting transmission in many foci in Africa more recently (4, 5, 15, 16), has led to high expectations that the global elimination of the disease using this therapeutic approach may be possible (1, 17). This shift in focus from reduction or control of the burden of the disease to transmission interruption has led to the ramping up of control activities using MDA (7, 17); however, it has also highlighted the difficulties of achieving the long-term interruption of parasite transmission in every infection setting by means of MDA alone (1, 18). This is particularly the case for populations that continue to remain exposed to high black fly populations and communities facing significant social and operational challenges in delivering the long-term high coverage MDA required for bringing about transmission elimination (1, 17, 19–21). These biosocial and technical challenges have led to the increasing recognition that cost-effective, socially acceptable, strategies that can complement MDA will need to be developed and assessed in order to successfully bring about the global elimination of the disease (22).

Recently, several workers have suggested integrating MDA with vector control (VC) to overcome these challenges (17, 23, 24). These workers stress how combining treatment of humans alongside the reduction of the prevailing vector density can serve to act on both microfilarial loads and vector-human contact rates synergistically in a population to facilitate a more rapid achievement of transmission elimination. One such recently investigated VC tool is the Esperanza window trap (EWT), first developed and used in Mexico, and subsequently evaluated in Burkina Faso and Uganda (1, 25–27), which has been shown to constitute a highly effective and easily implementable method for trapping *S. damnosum* black flies (25–28). The trap, utilizing a combination of olfactory cues and visual attractants to lure adult host-seeking blackflies, was initially used as a tool for xenomonitoring (21, 25, 29), but more recent studies have indicated that EWTs may also represent a portent means to reduce the local fly population potentially to levels that may facilitate the breakage of transmission (25, 28). Thus, EWTs baited with human sweat compounds and CO2 and deployed in households and schools were shown to be able to reduce *S. onhraceum* s.L. by 14-51% in Mexico (26, 30), while field trials carried out using traps baited with sweat-impregnated articles of clothing within classrooms in Northern Uganda showed that these could reduce *S. damnosum* biting rates in classes by as high as 91% (31). However, while these results highlight the utility of using EWTs as an effective VC tool particularly for locations where people congregate for carrying out daily activities, the evidence for their effectiveness in reducing fly numbers when deployed in outdoor settings has been variable (30–32). Moreover, there are also indications that the efficacy of these traps for trapping the same fly species could vary from one country to another (30, 31). These variable experimental results indicate that while EWTs may offer a promising tool for achieving black fly control, there is a need to evaluate the sources of the differences observed in fly trapping effectiveness under diverse transmission conditions if they are to be deployed successfully for assisting with eliminating the transmission of onchocerciasis.

Three sets of key questions require addressing for assessing the usefulness of EWTs, and indeed attractant- based insect traps in general, for reducing and breaking community-wide vector-borne macroparasitic disease transmission. The first are questions related to the components of an attractant insect trap network for effectively trapping and killing the target insect over a given landscape. These specifically include quantifying the impacts of trap densities, their placements, and attractiveness as well as effectiveness as a whole for capturing and killing insects in a given environment (33). The second set of questions is concerned with quantitative methods for relating recorded EWT fly collections in a locality to the corresponding local community vector biting rate, and to both vector and human infection incidences or prevalences. Addressing this association will be key to evaluating the utility of the EWT as a tool for bringing about community-wide parasite transmission elimination (34). Finally, assessment of the effectiveness of the tool for facilitating community transmission elimination also requires quantifying how combining EWT with MDA programs can have a synergistic effect in accelerating and sustaining the elimination of *O. volvulus* transmission in settings exposed to different black fly intensities or endemicity.

We have recently shown how simulation or computational models of macroparasite transmission can offer important quantitative tools for investigating the above questions by virtue of their ability to relate the components and actions of a control technology with the diverse states and processes governing the transmission dynamics of a disease in a host population (34–36). Indeed, the further capability of modeling frameworks for being able to integrate evidence from trial observations of a control measure in selected locations with the general process-based information contained within a theoretical model also means that such data-driven process-based simulation models can not only recapitulate existing empirical measurements but also predict the effects of the intervention under transmission conditions that have not yet been tested (37, 38). This means that such quantitative tools can offer a key scientific means for exploring the full range of possible transmission effects arising from an intervention via facilitating the effective extrapolation of existing trial observations to all relevant spatial domains or locations of interest (34). This is an important feature as it means that these modeling systems can afford generalizable or systematic insights into the operation of an intervention, which will be difficult to achieve using empirical field trials alone. This critical ability of models for being able to simulate the likely future states of an infection system undergoing a particular intervention in diverse settings has been key to their rising acceptance and adoption by decision makers as a means for evaluating intervention options that may best bring about the achievement of national and global disease management goals (17, 24, 35, 39–42).

Here, we describe the development and use of a data-driven process-based modeling framework for investigating the utility of combining EWTs with MDA as a new two-pronged strategy for progressing onchocerciasis elimination under typical endemic conditions. We note that according to WHO guidelines (1, 43), the decision for declaring that the ultimate elimination of parasite transmission has occurred in a setting is to be based on human and entomological infection surveys which will be carried out in three phases – with the first phase focused on assessing if specific WHO-defined human infection and infective thresholds in black fly populations below which transmission is thought not possible have been met, followed by additional entomological surveys in phase 2 (termed as the post-treatment phase (PTS)) and subsequent primarily parasitological surveys in phase 3 (the so called ‘post-elimination survey”) for confirming the absence of either ongoing, residual or resurging infections in vector and human populations respectively. As previously (34), we use the community-wide human infection threshold of 1% microfilarial (Mf) prevalence and an ATP (annual transmission potential) value of 20 infective bites per person per year defined originally as transmission elimination targets by the WHO for onchocerciasis in the foregoing analyses (44). We focus on the parameters for the field deployment of EWT plus MDA for enhancing each phase of the WHO transmission interruption assessment strategy in the analysis, with achievement of phases 2 and 3 examined by addressing the ability of the tool for bringing about long-term breakage of transmission post stoppage of MDA.

Our simulation system for addressing the utility of EWT plus MDA for accelerating and sustaining onchocerciasis transmission elimination has three major components. First, models of black fly trapping efficacies using various EWT field or network configurations are constructed. These are then combined with our population-level model describing the seasonal population dynamics of onchocerciasis. Finally, the coupled system is integrated with steps that include the estimation of initial conditions, launching of simulations, and carrying out analyses of model solutions for evaluating the impact of the supplementary EWT strategy for enhancing the efforts of MDA alone in meeting the infection and transmission elimination thresholds described above (45). We begin by presenting the mathematical basis of our capture probability model for EWTs based on the concept of an effective black fly attraction range and killing efficiency, and various parameters related to the deployment of insect attractant traps in a landscape, such as inter-trap distance, numbers of traps, areal coverage, size of area, and decay of trapping effectiveness (28, 31). We then describe how we couple this landscape-based spatially-explicit trap model with novel extensions of our seasonal population-level model of onchocerciasis transmission to allow simulations of the impact of using various trap configurations for reducing the community-level ATP, and by acting synergistically with MDA for reducing human infection to below the 1% Mf prevalence threshold. We performed these analyses by using transmission models calibrated to site-specific data from four previously described representative onchocerciasis endemic study villages from Uganda (34), which also allowed an examination of the impacts of seasonality and endemicity on transmission outcomes due to the EWT-based interventions. We end by discussing our results in terms of the potential of using EWTs acting in conjunction with MDA for achieving area-wide onchocerciasis elimination. We also highlight how population-level macroparasite transmission models, such as our deterministic onchocerciasis transmission dynamics model, can be effectively extended to address the spatio-temporal complexities that may govern the effectiveness of the EWT tool for bringing about parasite interruption in the real-world. Finally, we touch upon the value of using process-based simulation systems, such as the one developed here, as a means for extending the evidence regarding an intervention obtained from field trials so that higher-order knowledge generalizable to diverse transmission conditions can be discovered and used effectively in policy making.

## Methods

### Simulation system overview

Our data-driven computational system for carrying out simulations of the effects of EWT combined with MDA versus using MDA alone for reducing and eliminating onchocerciasis transmission at its core comprises the integration of three components (Figure 1). These include firstly the setting up and coupling of mathematical models of black fly capture probabilities over a landscape given various EWT trap configurations and parameters, including different trapping network patterns, trap attractive range, fly killing efficiency, and decay of trap attractiveness (31, 33), with our age structured-seasonal model of onchocerciasis transmission (34) that allows for the incorporation of both MDA and VC using EWT (see below). The parameters of the onchocerciasis model are then identified for a setting in this system via a calibration step, wherein we use our previously developed Monte-Carlo based Bayesian Melding (BM) data assimilation methodology to discover an ensemble of models that best-fit baseline site-specific infection data (35, 40, 46). Once these models are identified for a location, we implement and simulate the effects of control arising from the deployment of different EWT trap configurations and MDA strategies by linking their respective effectiveness in reducing the community-wide vector biting rate (EWT traps) as well as infection in the human population (MDA) on the overall dynamics of *O. volvulus* transmission in step three (31, 34). More specifically, here, we perform a comparative analysis of the impacts of using MDA alone versus combined MDA and EWT by evaluating: 1) the timelines predicted for each of these interventions to cross the specified community-wide Mf prevalence (=1%) and ATP (=20) elimination thresholds, and 2) their subsequent ability for bringing about long-term transmission interruption post-stoppage of MDA.

**Figure 1.**
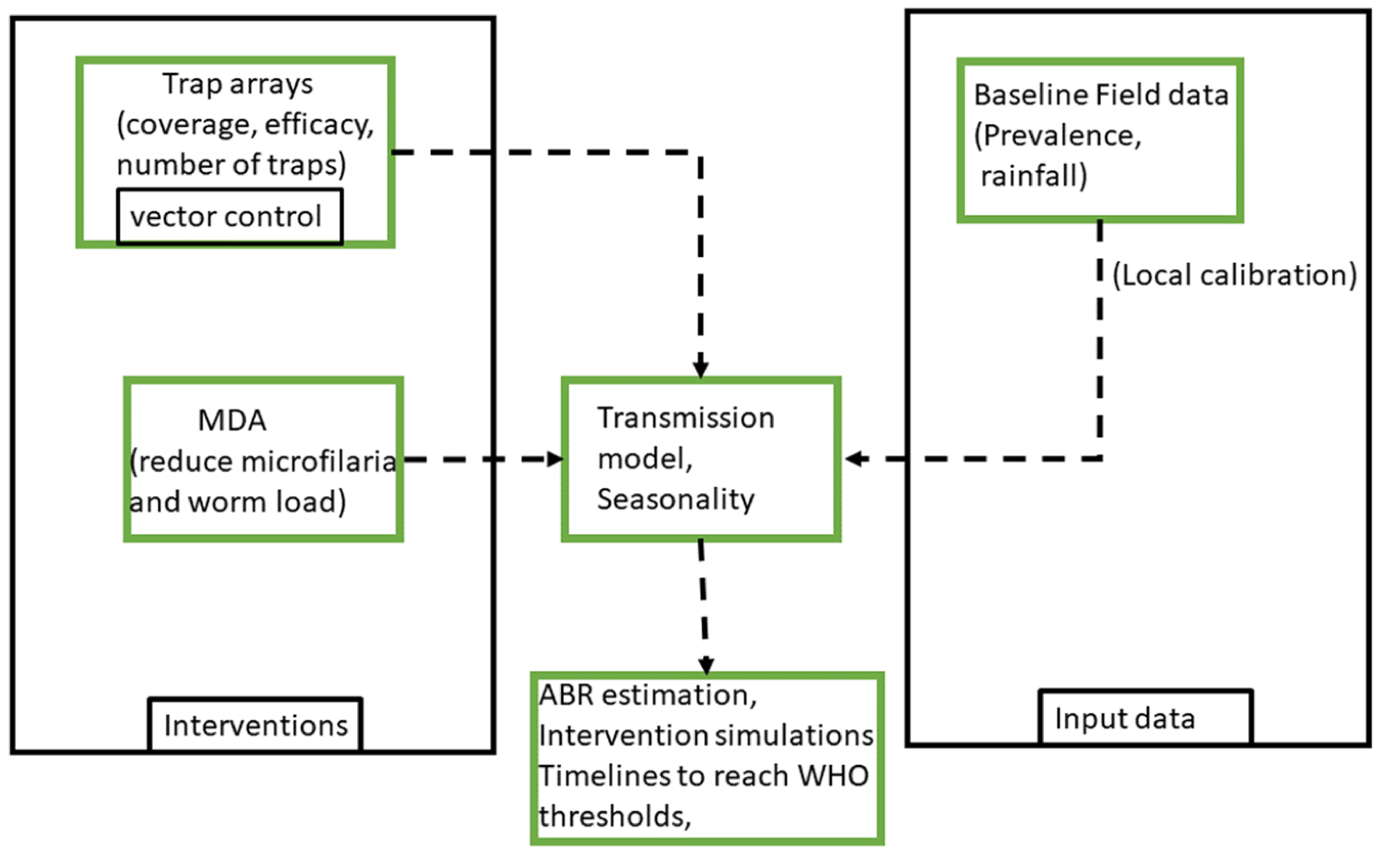
The computational framework for integrating interventions (EWT vector control and MDA interventions), the seasonal age-structured transmission model of onchocerciasis, and baseline field data.

### The mathematical model for quantifying EWT fly capture probability over an area

We begin by assuming that each EWT represents a square of dimension of *l* × *l* (Figure 2(a)). Typically, an array of such traps is placed at the village/site of interest; a possible array of traps is shown in Figure 2(b) assuming that the village is a rectangular box with an area *A*. The inter-trap distance is *d* while each individual trap in the array has a capture or attractant radius *r*. The capture radius defines a circular area around a trap where all the flies are trapped if they fly within this radius. If there are *Nt* number of traps, then the areal coverage *C_Tr_* in such a trap configuration or network can be defined as:

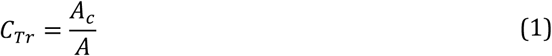

**Figure 2.**
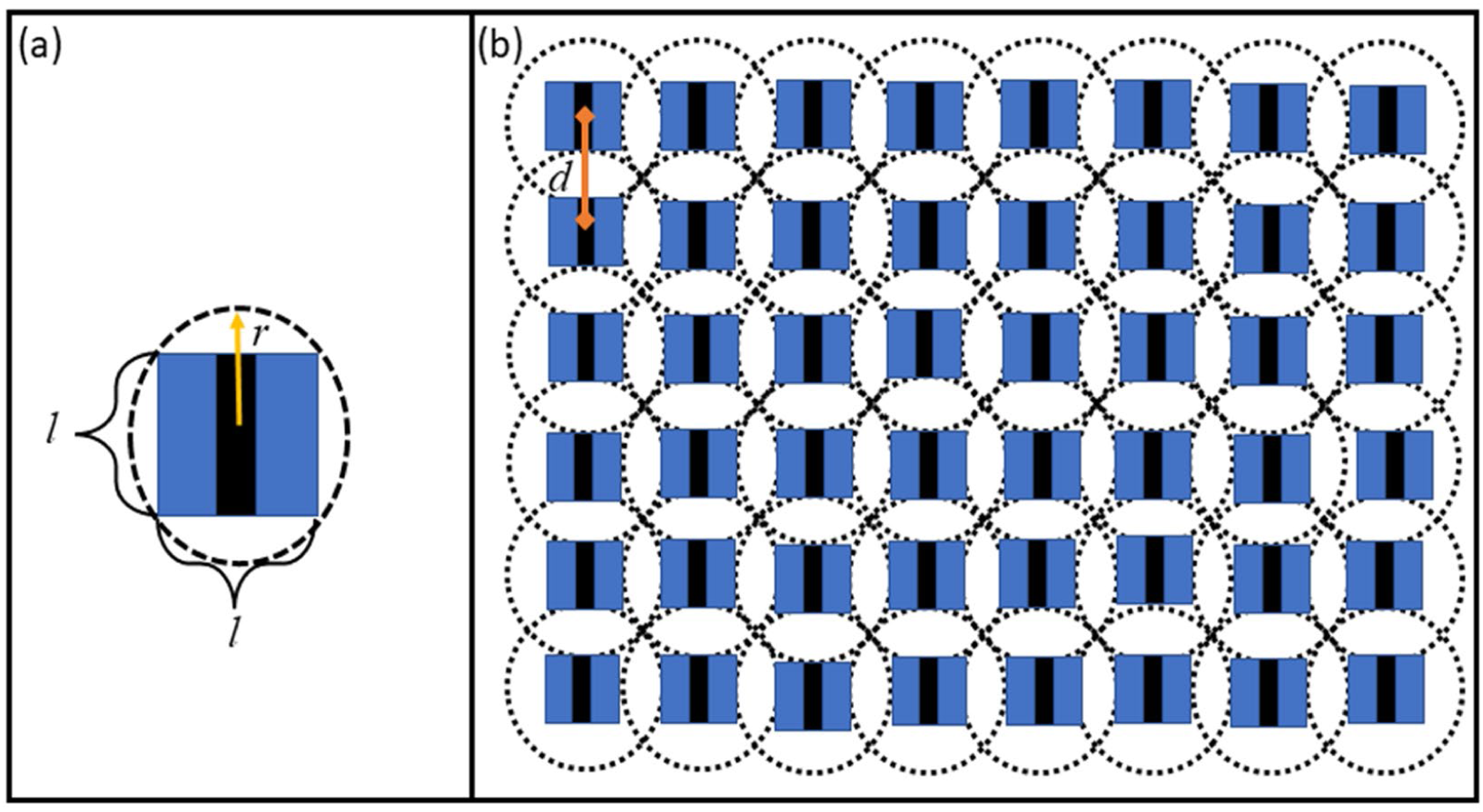
(a) An EWT trap with capture radius *r* and dimensions *l* × *l*. (b) A rectangular array of EWT traps placed over a landscape with a separation distance *d*.

where *A_c_* = *N_T_*π*r*^2^ is the combined capture area of all the traps. The total number of traps *N_T_* is then related to capture radius of individual traps *r* and the inter-trap distance *d* as follows:

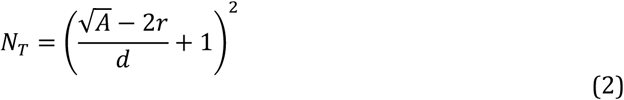

The above equations assume that the capture probability of a trap is unity within the capture or fly attractive radius of the trap and zero otherwise; this in turn can be used to determine the optimal distance *d* between traps to achieve a desired number of traps given a particular coverage, *C*_T_. When the capture probability of a trap is a continuous function of trap distance (instead of the Heaviside function considered here), the computation of capture probabilities via an array of traps becomes cumbersome as the capture probabilities of individual traps overlap. On the other hand, when fly capture by an individual trap is modeled by a hard radius of capture probability as described above, this approximation can be used to provide estimations of the upper limits on the number of traps required to achieve a given coverage as shown by Equation (2).

### Heterogeneous fly distribution

The above capture equations (Equation (1) and Equation (2)) represent the situation where the black fly population is homogeneously distributed and is assumed to bite hosts uniformly across a setting. In reality, fly biting and population distribution are likely to be spatially heterogeneous (47, 48). This spatial heterogeneity can govern the optimal number of traps required to achieve the desired coverage levels. To calculate the number of traps in a spatially heterogenous setting, we ideally require knowledge of the exact distribution of a fly population in such locations. However, the use of a discrete distribution, such as the Dirac-delta function to represent the localization of flies within an area of interest, wherein we assume that there are discreet areas in a setting with flies and areas without any flies (49), can allow the effective calculation of the number of traps required under this simplifying condition. Thus, assuming that the total area of sites where the flies are absent is *A*_Δ_, the coverage Equation (1) and the number of traps Equation (2) can be modified to obtain an areal trap coverage under heterogeneous fly biting, *C_Thet_* as:

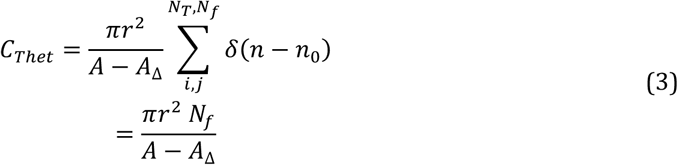

where *N_f_* is the number of sites where flies are present, *N*_T_ is the maximum number traps that can be accommodated for an inter trap distance *d*, and δ denotes the Dirac-delta function for locating the presence of flies at a specific point, *n*_0_, over a grid of discrete locations, *n*, within an area (ie. flies are present if *n* = *n*_0_ or absent otherwise) (49). In this formulation, it can be seen that if the trap coverage is set to be the same as in the homogeneous case the effective number of traps is simply equal to the number of sites where flies are present:

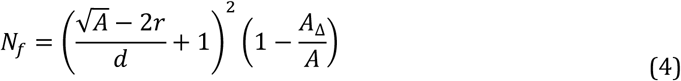

Equation 4 implies that the required number of traps decreases linearly with *A*_Δ_/*A*, where *A*_Δ_/*A* is a measure of the spatial heterogeneity observed for the biting black fly population.

To demonstrate how Equation 4 could be used in a field setting, we consider toy examples of fly heterogeneity and the corresponding optimal trap placement inside a village represented by a rectangular box as shown in Figure 3. In Figure 3(a) traps are placed by ignoring the fact that flies are concentrated at the periphery of the village (darker pixels), while in Figure 3(b) traps are placed in just those areas where flies are present. In this case, it is clear that accounting for fly heterogeneity would require a lower number of traps than otherwise to achieve a given coverage. The situation demonstrated in Figure 3(b) is similar to the one studied in reference (31), where the traps were able to reduce bites if placed in the periphery of fields, close to fly habitats. If traps are deployed by ignoring heterogeneity in flies as in Figure 3(a), then some traps among the array might be ineffective/irrelevant in reducing the number of bites. The number of ineffective traps in that case is given by the *N_ineffective_*=*N_T_* − *N_f_*:

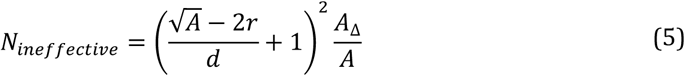

**Figure 3.**
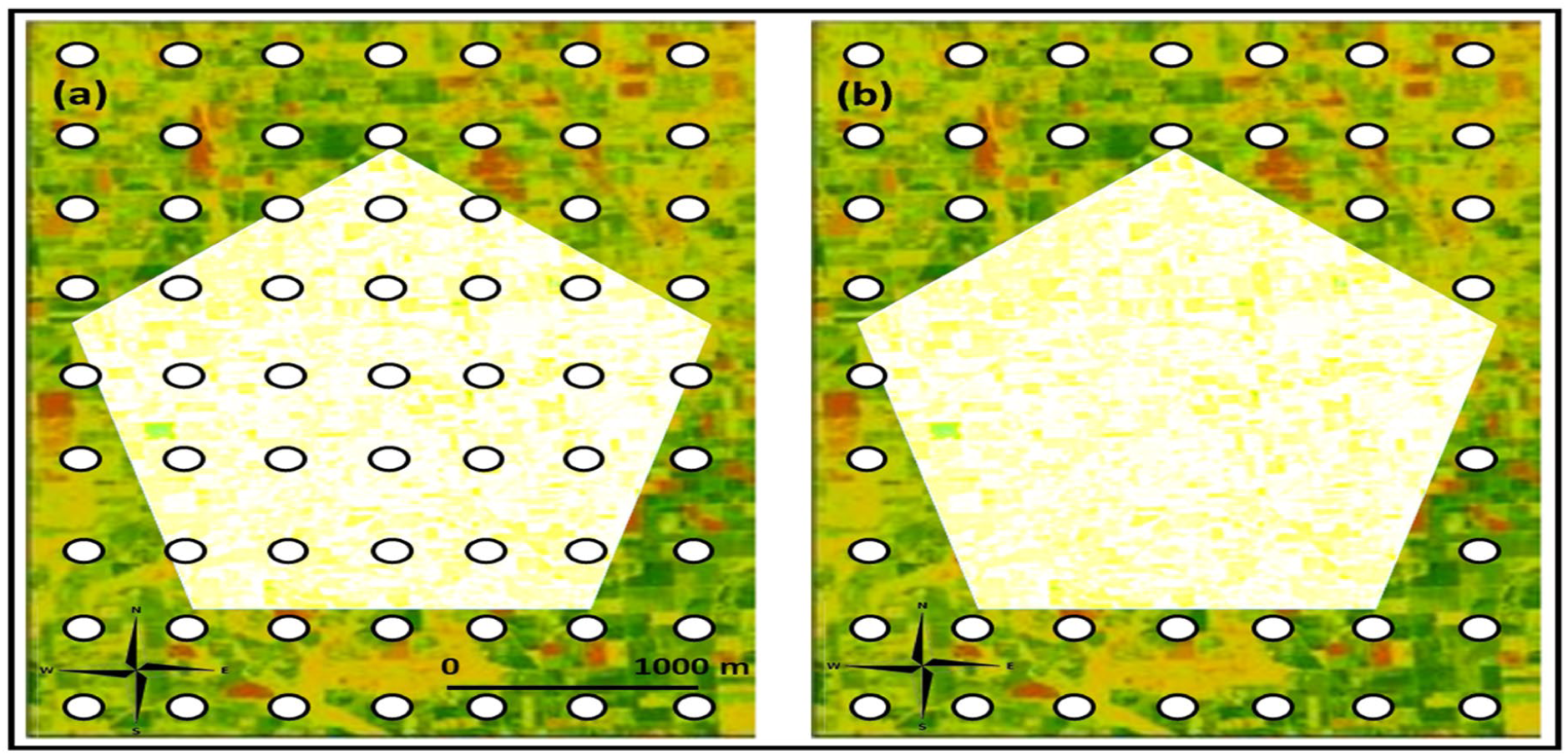
An array of EWT traps placed to capture flies in an heterogenous fly environment to achieve a desired coverage. We assume that flies are concentrated at the periphery of the shown region. (a) Placing traps flies by ignoring the fly distribution pattern. (b) Placing traps in the periphery. Both these configuration of trap networks achieve the same coverage but accounting for heterogeneity requires a lower number of traps.

Equation (5) shows that in contrast to *N_f_*, ineffective traps will increase linearly with the fly heterogeneity parameter.

### Onchocerciasis transmission model

The onchocerciasis transmission model used in this study is an immigration-death deterministic model describing the aggregate-level transmission dynamics of the parasite in both human and black fly populations. As detailed previously (34, 46, 50), the model is based on a set of coupled partial and ordinary differential equations, where the population-level age-structured pre-patent *P*(*a*, *t*) and patent *W*(*a*, *t*) worm burdens, microfilariae counts *M*(*a*, *t*), and acquired immunity (*I*(*a*, *t*)) in the human host are dynamically modelled by partial differential equations over time (*t*) and age (*a*), whereas the dynamics of infective L3 stage larvae per black fly (*L*) are modelled by an ordinary differential equation over time. The governing equations describing the dynamics of these system variables are:

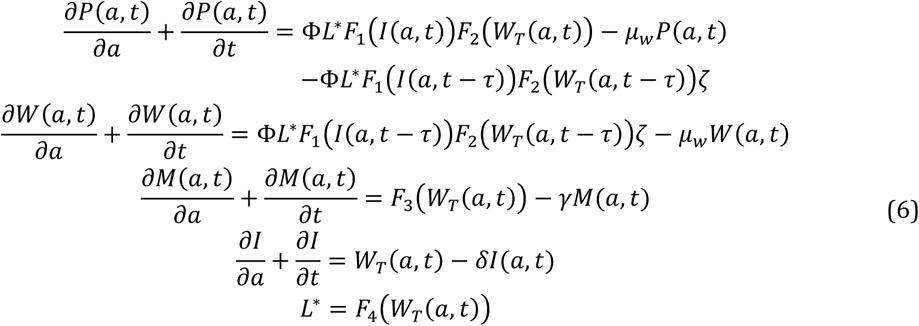

Here, the functions *F_x_* denote the density dependent mechanisms that govern the various transition or development rates of different states in the parasite life cycle (34, 46, 50). Note that some of these functions are dependent on the total worm load (WT = W(*a,t*) + P(*a,t*)), while the rest depend on the larval state (L*) and host immunity (*I*). Due to the significantly faster time scale of the infection dynamics in black flies compared to worm infection in humans, we further assume that the density of infective stage larvae in the vector population reaches a dynamic equilibrium (*L*^∗^) rapidly and use this simplification to simulate the infective stage larval dynamics. Details of all the parameters and functional forms are explained and described in the supplementary document Tables S1, S2.

### Bayesian Melding (BM)-based calibration of the model

We estimated parameters for the onchocerciasis transmission model via calibration of the model to a set of four previously published baseline infection datasets that spanned the range of endemic conditions that typically underlie the transmission of the disease (34) (Table S3). The model calibration exercise to identify best-fitting models for these locations was carried out using the flexible Monte-Carlo based BM data assimilation framework (35, 51). This method uses the following workflow. First, using the known ranges of the model parameter values, we define uniform prior distributions for each parameter and randomly sample with replacement from these distributions to generate *N* = 200,000 parameter vectors. The model is then run with each of the *N* parameter vectors, which generate *N* outputs predicting Mf prevalence by age. The simulated Mf prevalences are then compared against the age-stratified Mf prevalence data in each setting by calculating binomial log-likelihoods for each parameter vector. Since the datasets used in this work do not include age-stratified Mf prevalence data, age infection profiles were obtained from the observed overall community prevalence by following the procedure presented in Smith et al (36). Next, a Sampling-Importance-Resampling (SIR) algorithm is used to sample *n* = 500 parameter vectors with replacement from the pool of *N* parameter vectors with probabilities proportional to their relative log-likelihood values. This step generates the *n* parameter vectors most likely to represent the observed data. These *n* posterior parameter vectors are then used to compute distributions of variables of interest from the fitted models pertinent to this study (viz. age-prevalence curves and infection trajectories in both the human and black fly vector populations following interventions). Note that this approach, which results in the estimation and use of transmission models calibrated to local conditions, also provides an opportunity for examining the impacts of the variable endemic conditions that are typically observed in the field on onchocerciasis control dynamics (34, 46, 50).

### Steps to simulate MDA and EWT-based vector control

#### Simulating MDA control

We aimed to compare the complementary impact of EWT when added to onchocerciasis ivermectin-based MDA interventions with the outcomes arising from the use of the MDA strategy alone. The impact of MDA is modelled by assuming that the drug instantaneously kills *ω* and *ε* fractions of the adult worms (both pre- patent (*P*) worm and patent (*W*) adult worms) and Mf (*M*) populations. Because of uncertainty in the efficacy of annual IVM against adult *O. volvulus* parasites, the efficacy parameter *ω* is varied from 0.05 to 0.3 (39, 41, 42, 52–57) in the simulations. The effective population sizes of worms and microfilariae stages after drug treatment are given by:

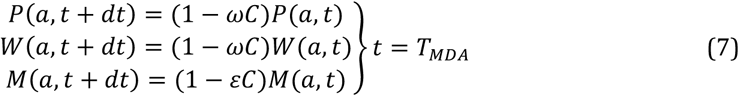

where *dt* represents a short time-period since T_*M*_, the time-point the *i^th^* round of MDA was administered, and the drug coverage at the population level is given by the parameter *C*. Each round of MDA kills a fraction of each worm stage (via values of *ω* and *ε*); however, the production of Mf by the surviving worms (53, 57) is reduced by a factor of (1- *δreduc*C):

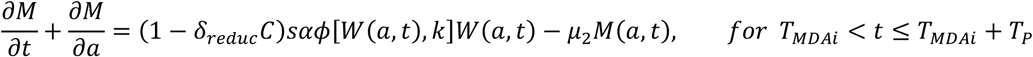

where *α* ′ = *α* (1 − *δ C*) is the reduced fecundity (over a period of *TP* months since the *i^th^* MDA) of the surviving adult worms at each MDA. The priors for *ω*, *ε* and T_*P*_ as given in Table S4 in the supplementary document.

#### Seasonality and simulating vector control using EWT

It has been shown that the use of EWT reduces the number of black fly bites on humans through trapping and killing of these flies when deployed persistently near fly habitats (31). We thus expect that the deployment of these traps across a locality will drive down the overall local black fly population and thereby decrease the aggregate vector-human biting or contact rate in the community (31). We further assume that one instance of EWT deployment constitutes placement of traps over a duration of a month with lures replenished on a monthly basis. On the other hand, seasonality in fly population dynamics, particularly associated with rainfall patterns, is a well-known driver of fly biting rates (22, 31). These factors suggest that simulating the impact of EWT-based vector control necessitates modeling the fly biting rates as a function of seasonal rainfall patterns (34), spatial biting patterns, and effects of deploying EWT traps over a setting on a monthly time- scale (31, 58). Here, we thus simulate the impact of EWT by first combining our previously developed model describing seasonally-varying black fly monthly infective biting rates (MTP) (34), with the vector control action of EWT in reducing the monthly fly biting rate, *MBR*(*m*), as follows:

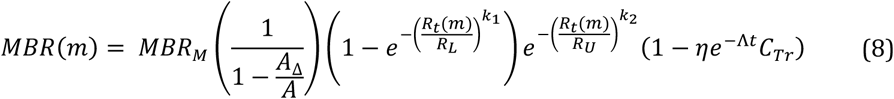

where *MBR_M_* is the maximum expected biting rate in a homogeneous environment, *R_U_* and *R_L_* represent upper and lower rainfall thresholds above and below which fly biting is greatly reduced, and *k_1_* and *k_2_* are shape parameters. Second, in environments with heterogeneous fly biting distribution, we scale up the mean maximum expected biting rate by a factor of 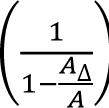 so that the average maximum expected biting rate over the area A of the village is still *MBR_M_*. Finally, the term (1 − η^*e−Λt*^*C_Tr_*) describes the reduction in MBR as a function of trap fly killing efficiency η, trap coverage, *C*_T_, as described in Equation 1 (the uniform biting case) or by Equation 3 (the heterogeneous biting case (*C*_Tℎ*ee*_)), and decay of trap fly killing effectiveness as defined by the term, *e*^−Λ*e*^. Note here in passing that this formulation also represents a novel method for incorporating the (discretized) variability in biting fly distributions that may occur in a landscape into a population-level parasite transmission model. The prior parameter ranges for η and Λ are given in Table S4 of the supplementary document. The other parameters in Equation 8 were obtained via the calibration of this rainfall dependent seasonal MBR model to monthly biting data from the control sites (no vector control) using BM with a pass/fail filter as described in (45).

### Modeling EWT and MDA intervention scenarios

We investigated the additional impact of combining EWT with MDA as a two-pronged integrated VC-drug treatment strategy for accelerating onchocerciasis transmission elimination by running our coupled EWT trap-transmission dynamics simulation model to mimic the effects of different control scenarios related to various EWT configurations and MDA strategies. These included modeling: 1) the impacts of various EWT configurations (variations in trap capture or attractant radius, fly killing efficiency, inter-trap distance, coverage and trap numbers) in reducing ATP, and 2) comparing annual and switch MDA (annual MDA for 10 years followed by a switch or change to bi-annual MDA) interventions alone versus including EWT into these drug regimens on the timelines required to cross target community-wide Mf (=1%) and ATP (=20) thresholds. Transmission models fitted to the four study sites used here were further employed to explore the impact of endemicity on the timelines to elimination arising from the implementation of the above intervention scenarios. Further, different EWT deployment frequencies were also simulated to investigate the impacts of using EWT a month before the peak fly biting season, for three months over the peak biting season and at monthly intervals during the year. This allowed examination of the impact of seasonality in the black fly population dynamics on the optimal timing of EWT deployment for reducing the ATP in a community. Note here that while our transmission model is calibrated to reductions in MBR based on empirical data from the EWT field experiments that specifically focused on the monthly effectiveness of the traps for reducing this variable (31), we predict the corresponding effect on ATP by multiplying the estimated MBRs (Equation 8) by the monthly population averaged L3* values, and then summing the resulting monthly transmission potentials (MTPs) to obtain the yearly ATP (34):

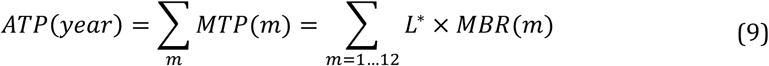

All the vector control scenarios in combination with MDA were modelled at the WHO-recommended drug coverage of 80%.

### Impact on long-term transmission elimination and recrudescence

We addressed this critical question by evaluating the behavior of the model trajectories once population infection prevalences in each of the four study sites are predicted to cross the threshold of 1% Mf following MDA, and all further drug applications are stopped. The impact of including EWT versus no vector control on elimination and recrudescence was compared post stoppage of MDA at two time points – five years post MDA (to cover the PTS assessment period) and fifteen years post MDA (to evaluate the longer-term impact of EWT on parasite elimination). Following Sharma et al.(59), we calculated the probability that transmission extinction has been achieved in a site due to an applied intervention by quantifying the proportion of the best-fit SIR model prevalences that were declining or declined to zero (ie. models giving rise to Mf prevalence curves with significant negative slopes) by the end of the 5 or 15-year simulation period following the crossing of the 1% Mf threshold value. The recrudescence probability for a given site was similarly calculated as the proportion of the total SIR selected model runs that managed to revive and generate positive increases in Mf prevalence (ie. give rise to Mf curves with significant positive slopes) by the end of each of the above simulation periods once MDAs are stopped. Finally, the curves having non-significant and fluctuating positive or negative slopes are considered as those exhibiting transient dynamical behavior (59).

## Results

### Identifying locality-specific seasonally-driven onchocerciasis transmission models

The impact of using EWT in the field in conjunction with annual and switch MDA on the timelines required to breach the community-wide 1% Mf prevalence and 20 ATP thresholds was investigated in this work using our data-driven seasonal MBR-based transmission model calibrated to four endemic village communities in Uganda (34). The fits of the model to baseline MF age-prevalences observed in each of these study sites using the BM-based data assimilation approach are shown in Figure S1 in the Supporting Information. Note as described previously (34, 35, 46), given uncertainty in parameter values, we identify 500 best-fitting models to characterize the local parasite transmission dynamics in a setting. The good fits of the models to data portrayed in Figure S1 amply highlight the capability and versality of the BM framework for reliably capturing and differentiating between the transmission dynamics of onchocerciasis that prevailed pre-intervention in the four settings.

### EWT parameters and the annual transmission potential (ATP)

We began our investigation of the impact of using EWT for eliminating parasite transmission from *Simulid* populations by evaluating the resulting village-wide reductions predicted for ATP as a function of differences in the parameters related to the deployment of traps (or the configuration of the trap network) in an endemic area. The results illustrated in Figure 4 are for the transmission model estimated for the village of Masaloa, and are based on assuming that the village encompasses a rectangular area of 1 *km*^2^, each trap has an fly killing efficacy of 70% (31) and a capture or attractant radius of 200m, and that traps are deployed on a monthly basis for a year. The simulations show firstly that decreasing the inter-trap distance, *d*, will decrease the ATP in this village, such that approximately at an inter-trap distance of 490 m (vertical solid line in Figure 4 (a)) the ATP threshold of 20 infective bites per person per year will be breached. Figure 4 (b) on the other hand highlights how changes in the inter-trap distance will affect the areal coverage, *C*_T_, required to keep the ATP below the threshold of 20 infective annual bites per person. Thus, while an inter-trap distance of approximately 600m will result in a coverage value of 50%, this will clearly be insufficient to breach the ATP threshold. By contrast, only the decrease of this distance to 490m will allow the crossing of this threshold as a result of the increase brought about in areal coverage to approximately 60%.

**Figure 4.**
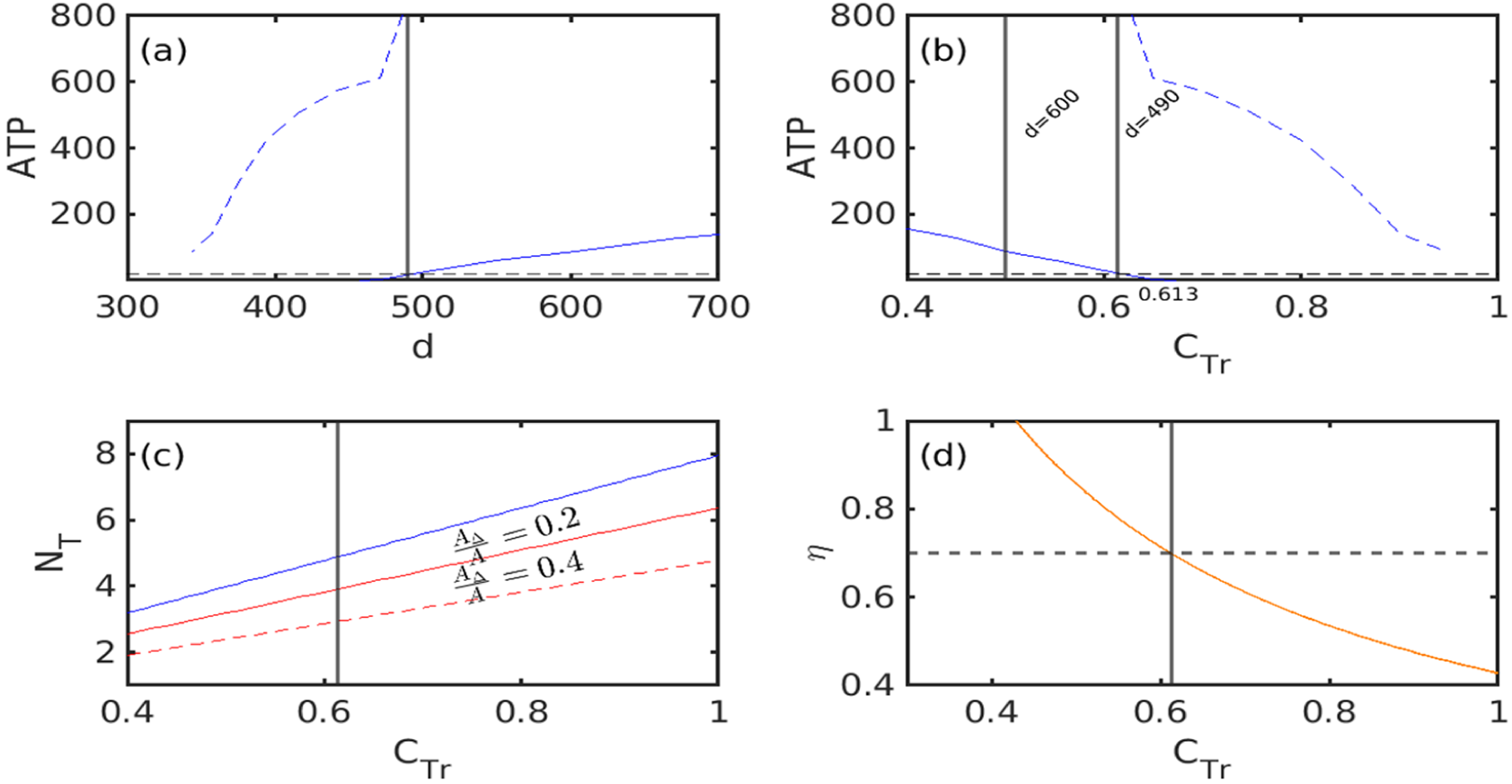
Curves in (a)-(b) show mean (solid blue) and 95 % confidence intervals (dashed blue) for ATP changes when an array of traps is deployed. The ATP decreases to the values as shown, starting from the baseline median value, after deployment of EWT for a year to reach the ATP threshold of 20 bites per year per person. Trap fly killing efficacy in these simulations is assumed to be 70 %. In (a), the lower inter-trap distances mean greater reduction in ATP, whereas in (b) higher coverage corresponds to greater reduction in ATP. The dashed- black horizontal line in (a) and (b) shows the ATP threshold of 20 bites per person per year. (c) The relationship between number of traps and the corresponding coverage achieved for a homogeneous fly distribution scenario (blue line) for two different values of the fly heterogeneity parameter 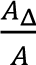 (solid-red 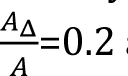 and dashed-red line 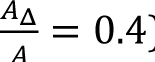). (d) The relationship between coverage and fly killing efficacy of traps, *η*, for attaining the 20 ATP threshold. The vertical black line in (a) - (d) show the values of inter-trap distance, coverage and number of traps required to cross the ATP threshold, while the horizontal dashed line in (d) corresponds to a trap fly killing efficacy of 70%, the value of efficacy used for (a)-(c). Importantly, the efficacy and coverage values in the region above the orange curve in (d) will result in the ATP threshold reached within a year. Each trap has a capture radius of 200 m and the study site for this figure is Masaloa.

The results in Figure 4 (c) and Table 1, however, indicate that the number of traps required to breach the 20 ATP threshold is determined by both trap coverage and the area of a village. Thus, Figure 4 (c) shows that given a trap fly killing efficacy of 70%, capture radius of 200m, an inter-trap distance of 490m and assuming that the area of Masaloa is 1 *km*^2^, the total number of traps required to achieve a coverage of 60 % (for breaching the ATP threshold in this example) is approximately 5 traps, all placed along a rectangular array as shown in Figure 3 for the homogeneous biting case (solid blue curve). The required number of traps for different coverages and village areal sizes along with corresponding inter-trap distances are further recorded in Table 1, which show that while the number of traps required to achieve 60-70% coverage will generally increase with area (for all fly distribution patterns), for a given areal size, this number will decline dramatically as the trapping radius and distance between traps increase. This three-way relationship indicates that as the capture radius of the trap increases, traps can be placed at greater between-trap distances, which in turn will lower the required number of traps for crossing a given ATP threshold.

**Table 1.** Relationship between number of traps, trapping radius, inter-trap distance, and heterogeneity in fly biting distribution accounting for size of a setting. Trap fly killing efficiency is 0.7-0.8. Results are from the use of the best-fitting onchocerciasis models for Masaloa.

| R<br>(trapping<br>radius,m) | d<br>(distance<br>between<br>traps,m) | $N_T$ (number of traps to achieve 60-70 percent coverage) | | | | | | | | | | | |
| --- | --- | --- | --- | --- | --- | --- | --- | --- | --- | --- | --- | --- | --- |
| | | Area<br>$A=1\text{ km}^2$ | | | | Area<br>$A=5\text{ km}^2$ | | | | Area<br>$A=10\text{ km}^2$ | | | |
| | | $\frac{A_\Delta}{A}$<br>= 0 | $\frac{A_\Delta}{A}$<br>= 0.2 | $\frac{A_\Delta}{A}$<br>= 0.4 | $\frac{A_\Delta}{A}$<br>= 0.8 | $\frac{A_\Delta}{A}$<br>= 0 | $\frac{A_\Delta}{A}$<br>= 0.2 | $\frac{A_\Delta}{A}$<br>= 0.4 | $\frac{A_\Delta}{A}$<br>= 0.8 | $\frac{A_\Delta}{A}$<br>= 0 | $\frac{A_\Delta}{A}$<br>= 0.2 | $\frac{A_\Delta}{A}$<br>= 0.4 | $\frac{A_\Delta}{A}$<br>= 0.8 |
| 50 | 115 – 124 | 69<br>– 78 | 55<br>– 63 | 41<br>– 47 | 14<br>– 16 | 1706<br>– 1950 | 1365 –<br>1560 | 1024 –<br>1169 | 342<br>– 390 | 6825<br>– 7800 | 5500<br>– 6235 | 4100<br>– 4680 | 1365<br>– 1560 |
| 100 | 235 – 256 | 18<br>– 20 | 14<br>– 16 | 11<br>– 12 | 3 – 4 | 427<br>– 487 | 342<br>– 390 | 256<br>– 293 | 86<br>– 98 | 1706<br>– 1950 | 1365<br>– 1560 | 1024 –<br>1169 | 342<br>– 390 |
| 200 | 490 – 564 | 5 | 4 | 3 | 1 | 107<br>– 122 | 86<br>– 98 | 64<br>– 74 | 22<br>– 25 | 427<br>– 487 | 342<br>– 390 | 256<br>– 293 | 86 – 98 |
| | | $N_T$ (number of traps to achieve 90-100 percent coverage) | | | | | | | | | | | |
| 50 | 87 – 93 | 115<br>– 128 | 92<br>– 102 | 69<br>– 77 | 23<br>– 26 | 2870<br>– 3200 | 2292<br>– 2550 | 1720<br>– 1910 | 573<br>– 637 | 11460<br>– 12735 | 9170<br>– 10190 | 6860<br>– 7640 | 2292<br>– 2550 |
| 100 | 172 –<br>183 | 29<br>– 32 | 23<br>– 26 | 18<br>– 20 | 6 – 7 | 717<br>– 796 | 573<br>– 637 | 430<br>– 478 | 144<br>– 160 | 2865<br>– 3184 | 2292<br>– 2550 | 1720<br>– 1910 | 573<br>– 637 |
| 200 | 330 –<br>360 | 7 – 8 | 6<br>– 7 | 4 – 5 | 1 – 2 | 180<br>– 200 | 144<br>– 160 | 108<br>– 120 | 35<br>– 40 | 717<br>– 796 | 573<br>– 637 | 430<br>– 478 | 144<br>– 160 |

Additionally, the optimal coverage (and hence number of traps) needed generally to cross the ATP threshold is also influenced by trap efficacy in killing the trapped flies. This relationship is shown in Figure 4 (d), which demonstrates that as fly killing efficacy, η, increases, the ATP threshold can be achieved at a lower coverage and vice-versa (as denoted by the orange curve). Importantly, the results also indicate that there is a critical trap fly killing efficacy (approximately 45% in the present example) below which the ATP threshold cannot be achieved within a year even when coverage is maximal (100%). By contrast, achieving any of the efficacy and coverage values above the orange curve will lead to the ATP threshold of 20 infective bites per person per year being reached within a year of monthly EWT deployment (Figure 4(d). These results are derived from the models fitted to the village of Masaloa. Similar results were obtained for the other three study villages investigated here, indicating the generality of these findings.

### Spatial heterogeneity in fly distribution, EWT coverage, and ATP

We used the ratio *A*_Δ_/*A* as a measure of spatial heterogeneity in the distribution of the fly population at a given site (total area with no flies *A*_Δ_ over total area *A*) to calculate the number of traps required to achieve a desired coverage under the heterogeneous fly biting scenario. Equation 3 implies that the number of traps to meet a given coverage decreases as the fly population heterogeneity *A*_Δ_/*A* parameter increases for a village with area *A*. This outcome can be seen in Figure 4(c) for two values of fly distribution heterogeneity parameter 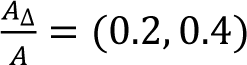 and assuming an area of 1 *km*^2^ for Masaloa. However, in general, the number of traps required to achieve the same coverage for a given trap fly killing efficiency is driven by values of *A*_Δ_ and *A*, the village size, and both the trapping or attractant radius and between trap distance. This can be seen from the results given in Table 1, which shows firstly that for a given mix of trapping radius and inter-trap distance, the number of traps a village requires to meet the same coverage will decrease as heterogeneity increases. Secondly, for a given fly distribution, the number of traps required to meet a particular coverage will also decrease with increasing trapping radius and inter-trap distance. On the other hand, while the number of traps required to achieve the higher coverage of 90-100% will be greater than for achieving the lower 60-70% coverage for all values of biting fly distributions, these can be placed at smaller inter-trap distances to attain the increased coverage for the same trap efficiency. Finally, these calculations also provide a guide as to the feasibility of using EWT as a vector control measure in different settings varying in areal size and fly distribution. Thus, while in smaller sized village settings, eg. in the case of the 1 or 5 *km^2^* areas studied here, the number of traps required to attain a coverage of 60-70% can be as low as 1 to 25 when the trapping radius and biting heterogeneity is highest (i.e. for 200m and 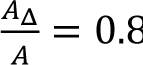), these can be as high as 86 to 98 for the same conditions and trap killing efficiency in the largest area (i.e. 10 *km*^2^) examined (Table 1). Although these predictions are for the simplified rectangular areal case, they do suggest that feasibility considerations might limit the use of traps alone in large areal settings, especially when trap capture radius is low and high coverages are required even in the case when fly biting heterogeneity is substantial (eg. for radius of 100m, 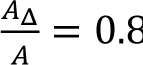, and village size of 10 *km*^2^, the number of traps to attain 90-100% coverage can be as high as 573-637 (Table 1)).

### Projected impacts of EWT and MDA on intervention durations for achieving parasite elimination

Next, the seasonal MBR-based transmission model was coupled with EWT field deployment configurations and either annual or switch MDA for simulating the timelines needed to reach the two elimination thresholds investigated. The baseline models identified for each of the 4 study villages were used to further evaluate different EWT temporal deployments, viz. traps placed monthly, before and during the peak biting season, used alongside either of the MDAs in these simulations. The area of the simulation village is assumed to be 1 *km*^2^ while the trap capture radius and fly killing efficiency is set to 200 m and 75%in all the simulations. Simulations were run for both the homogeneous versus heterogeneous fly biting scenarios. All the simulations were also carried out using a MDA coverage of 80%, while the EWT coverage was varied. Note also that given we identify 500 models to provide an adequate representation of the parasite transmission dynamics in a locality, the intervention durations predicted for an intervention are based on when 95% of these models cross the 1% Mf and ATP (=20) thresholds respectively in each study site.

Figures 5 and 6 show the durations in years required to cross the Mf and ATP elimination thresholds for annual and switch MDA without (the base case) and with EWT deployed at coverages ranging from 60% - 70% predicted for the village of Masaloa under the homogeneous fly biting scenario – predictions of the timelines for all four sites for both these MDA regimes alongside various EWT coverages are summarised in Tables S5 – S9. The number of years of MDA required to cross the thresholds for Masaloa with MDA alone were found to be as follows: ATP is reached by an average of 12 years with annual MDA, whereas it takes a mean of 11 years with switch MDA (Figures 5 and 6; Table S5). The corresponding timelines to breach the Mf threshold are, however, 20 and 15 mean years for each of these MDA regimens (compare Figure 5 (a) versus 6 (a); Table S5). On the other hand, when EWT is deployed monthly together with annual and switch MDA, the ATP threshold for this village is predicted to be reached in as little as a mean of just 1 year, while the time to reach the corresponding 1% Mf threshold is reduced to means of 18 and 14 years (Figures 5(b) versus 6(b); Table S5). Interestingly, while the frequency of using EWT (i.e., a month before peak fly biting season, during the three months of the peak biting season, and monthly throughout the year) had a significant impact in reducing the timelines in years to breach the ATP threshold for each village, with most savings obtained for monthly deployment and least in the case of usage just before the peak biting season, this factor did not have an effect on the years saved with respect to crossing below the Mf threshold (Table S5). Overall, however, the results indicate that a significant number of intervention years will be saved when EWT is used alongside MDAs (as opposed to MDA alone), with savings in mean years achieved ranging from 1-17 years in the case of combined annual MDA (80% coverage)-EWT (60-70% coverage) compared to 1 to 12 years when switch MDA is applied with EWT at the same coverage levels (Table S5). Reducing average EWT coverage (from 65% to 45%) and hence number of traps in each village will, as expected, significantly reduce these savings in intervention years (and conversely increase timelines to reaching either threshold), although again this impact will primarily be seen for the 20 ATP rather than the 1% Mf threshold (Tables S6 – S9).

**Figure 5.**
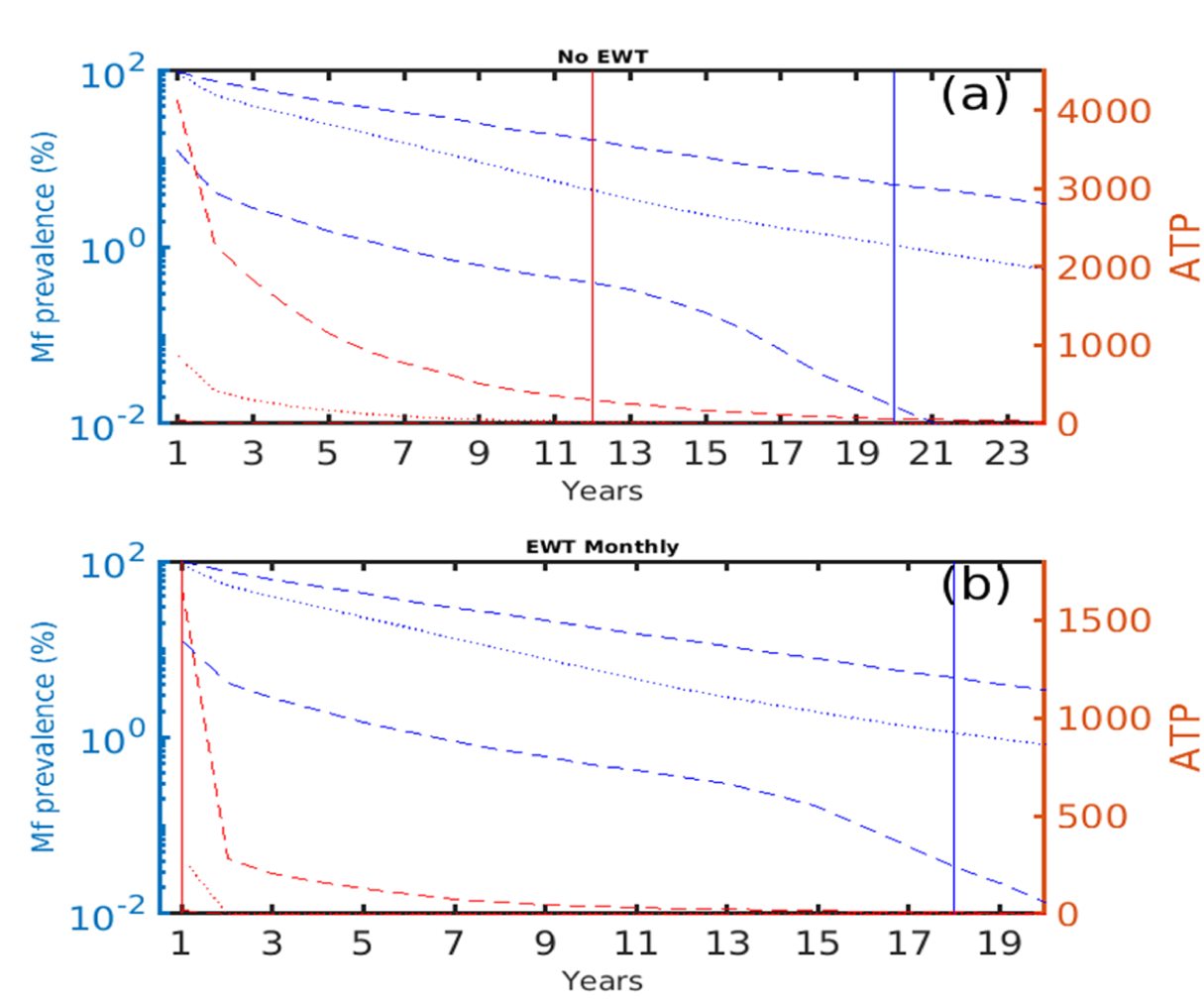
Mean (solid curves) and 95 % confidence intervals (dashed curves) of Mf prevalence (blue) and annual transmission potential-ATP (red) in the study site Masaloa as a function of time when: (a) only annual MDA and (b) annual MDA and monthly EWT are used in the homogeneous fly biting scenario. The times to reach the two thresholds (vertical red (20 ATP) and blue (1% MF prevalence)) are significantly reduced by the use of annual MDA and monthly EWT as compared to MDA alone. Here the EWT coverage is 60% to 70% assuming a trapping radius of 200 meters. The fly killing efficiency of the traps is assumed to be 70% to 80%.

**Figure 6.**
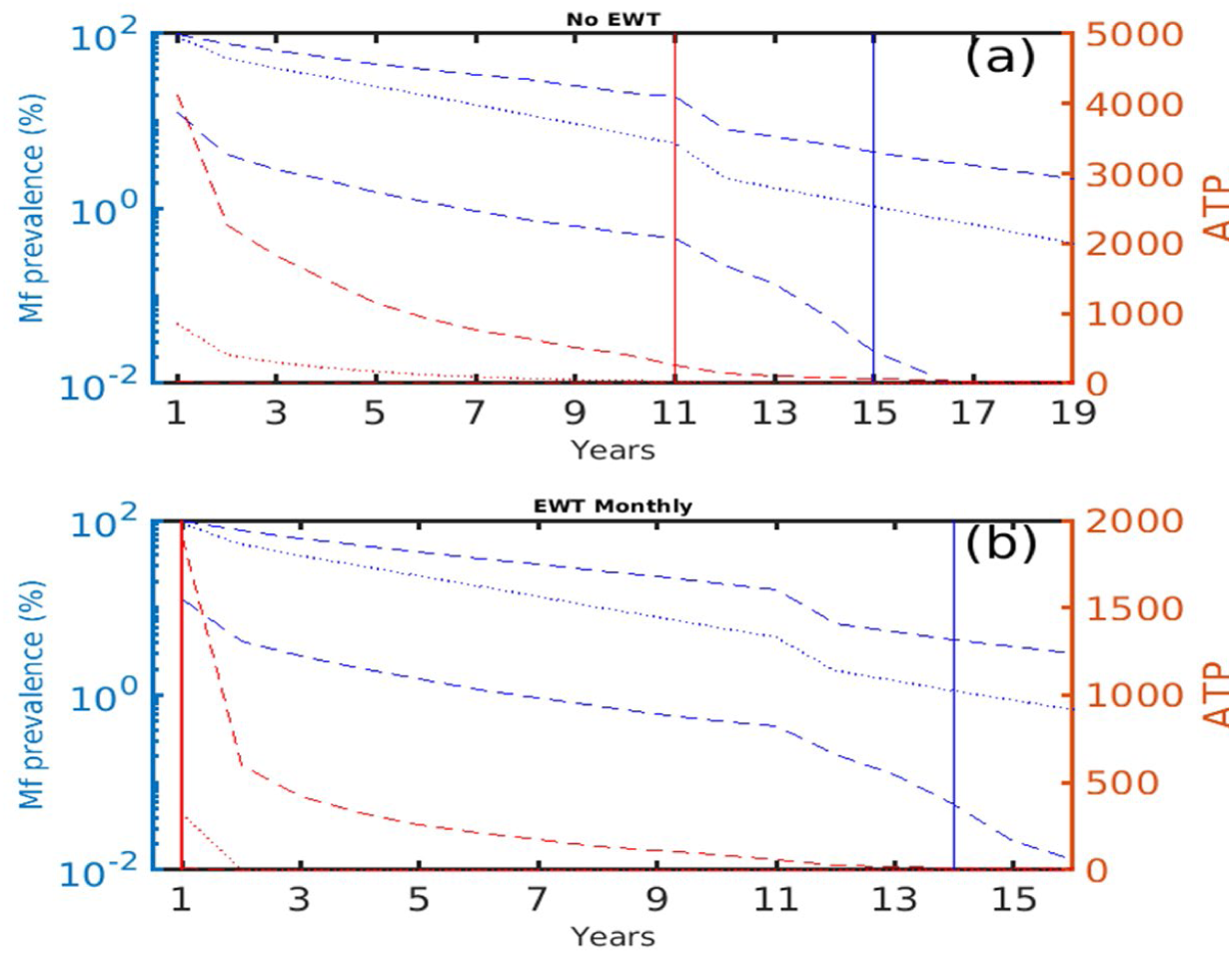
Mean (solid curves) and 95 % confidence intervals (dashed curves) of Mf prevalence (blue) and annual transmission potential-ATP (red) in the study site Masaloa as a function of time when: (a) only switch MDA (annual MDA for 10 years followed by bi-annual MDA) is used, and (b) when switch MDA regime is used in tandem with monthly EWT in the homogeneous fly biting scenario. The times to reach the two thresholds (vertical red line (20 ATP) and blue line (1% MF prevalence)) are significantly reduced as compared to MDA alone. Here the EWT coverage is 60% to 70% assuming a trapping radius of 200 meters. The fly killing efficiency of the traps is assumed to be 70% to 80%.

### The impact of heterogeneous fly distribution on timelines to elimination

In the scenario where fly biting is heterogeneously distributed, such as occurring only in the periphery of a village, the simulations indicate that paradoxically the times required to reach the community-level 1% Mf and 20 ATP thresholds will be significantly higher than those predicted for the homogeneous setting where the average biting rate is uniform across the village (Figure 7). Indeed, this simplified approach for incorporating fly biting heterogeneity reveals a steeply rising nonlinear relationship between the parameter 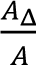 and the times to reach both the thresholds. This relationship can be observed from the results displayed in Figure 7 when EWT at fly killing efficacy ranging between 70 to 80% is deployed monthly at a coverage of 60 to 70%.

**Figure 7.**
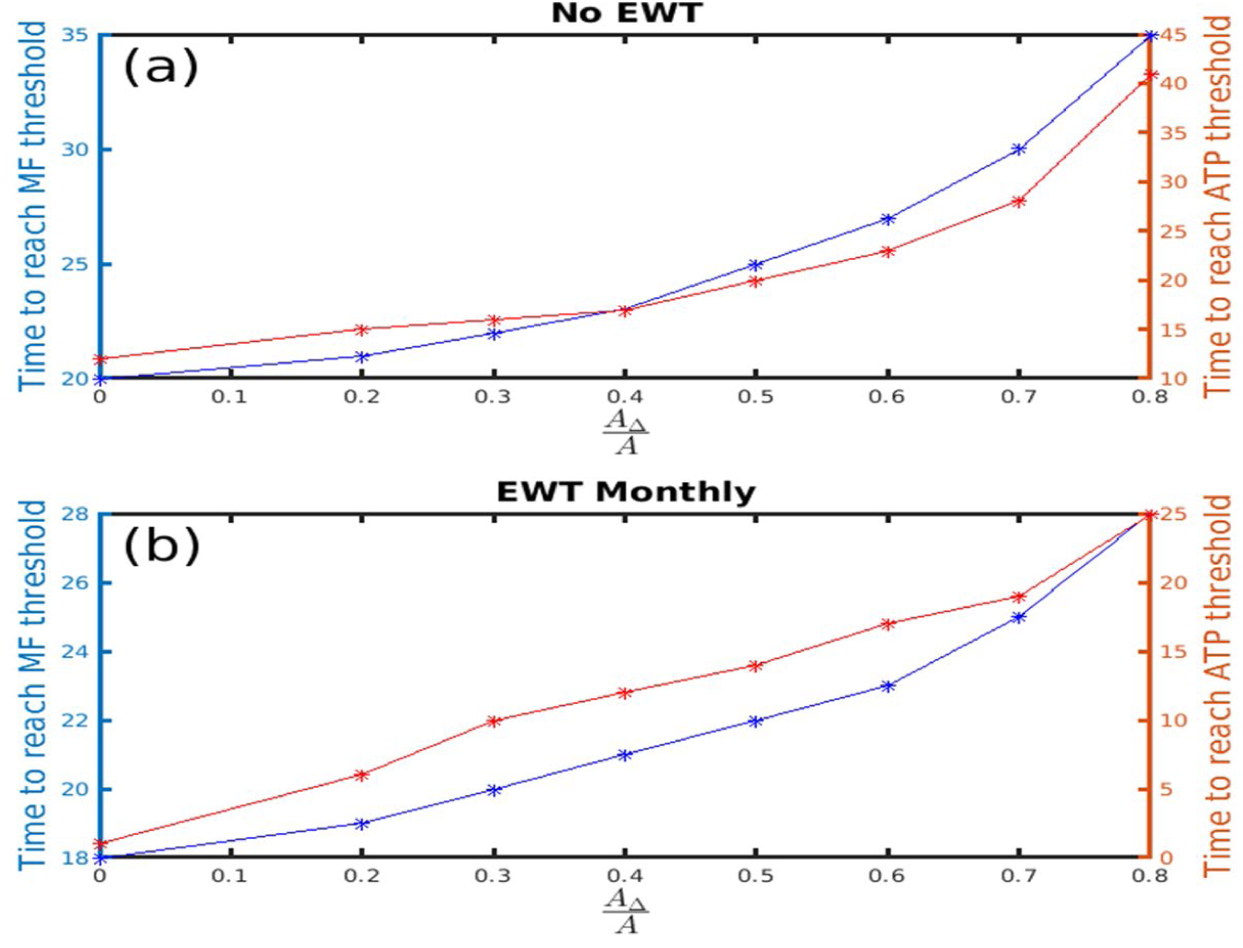
The time to reach the Mf and ATP thresholds as a function heterogeneous biting. Both (a) and (b) show the time to reach Mf elimination (blue curve) and ATP (red curve) thresholds in the study site of Masaloa as a function of time when: (a) annual MDA alone is used, and (b) when annual MDA is used in tandem with monthly EWT. The times to reach the WHO set thresholds increase nonlinearly as biting heterogeneity increases. Here the EWT coverage is 60% to 70% assuming a trapping radius of 200 meters. The fly killing efficacy of traps is 70% to 80%.

Figure 8 further depicts the number of years saved by the use of combined EWT and annual MDA compared to when yearly MDA alone is used in relation to variations in the fly biting heterogeneity and trap coverage. The results show, firstly, that combining EWT with MDA under both the homogeneous and heterogeneous fly distribution scenarios will result in significant numbers of years saved for breaching both thresholds in comparison with using MDA only (Kruswall − Wallis test comparing timelines to the Mf and ATP thresholds for each of the EWT/MDA scenarios shown in Figure 8 versus MDA alone: p <0.05 in each case). For the homogeneous scenario, the mean number of intervention years saved can range from between 7 to 10 years depending on trap coverage and the threshold target (whether Mf or ATP). By contrast, this can reach as high as between 10 to 40 years in the case of the heterogeneous fly biting scenario.

**Figure 8.**
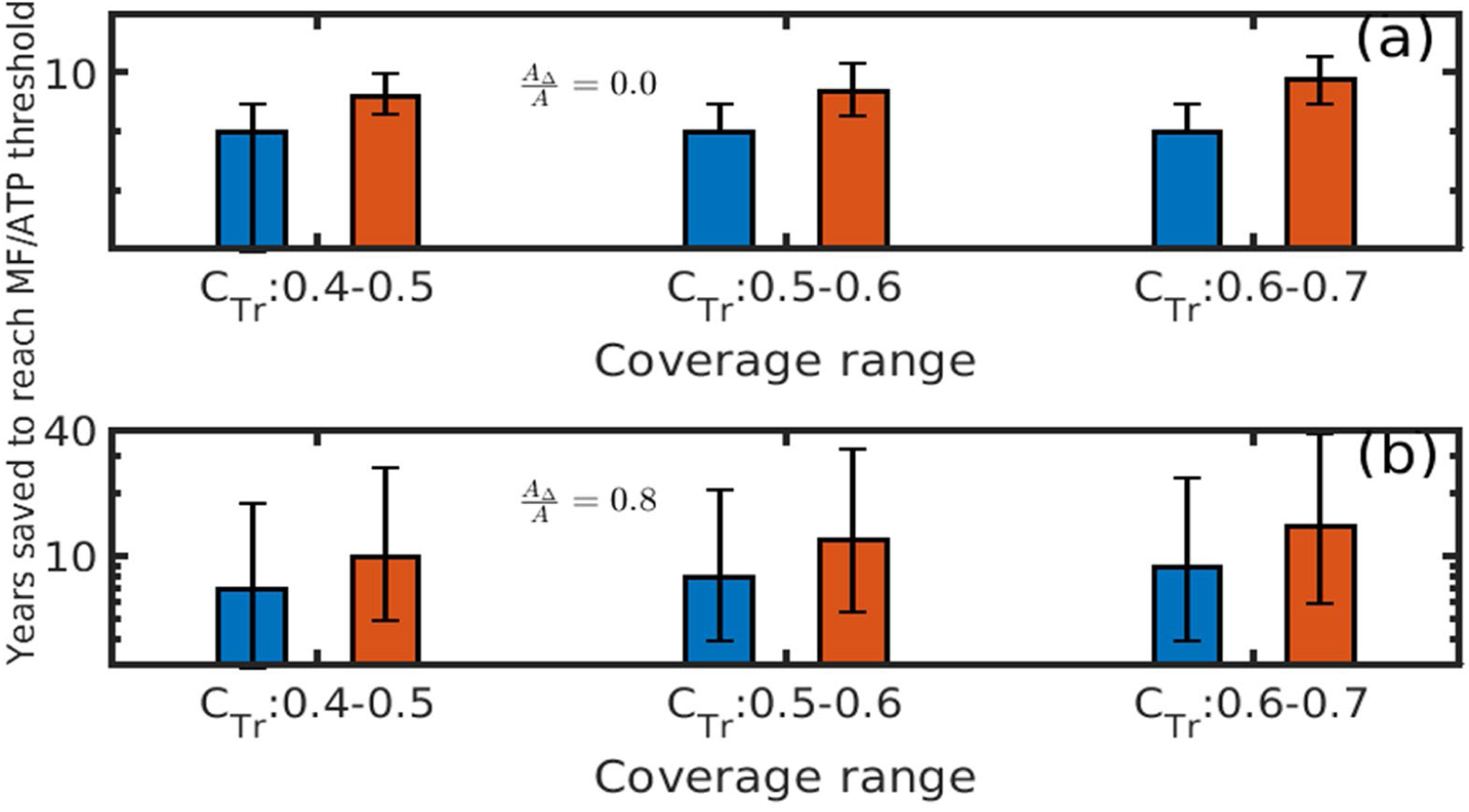
Bar plots indicating years saved in reaching the ATP (red) and Mf (blue) thresholds when EWT is deployed monthly alongside annual MDA in comparison with the use of annual MDA alone for increasing coverage values in the (a) homogenous fly biting case and (b) for the heterogeneity factor 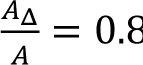. The results in shows that the combined use of traps and yearly MDA increases the number of years saved in reaching Mf and ATP thresholds for a given coverage compared to when using MDA only. For the homogeneous scenario in (a), the mean number of intervention years saved range from between 7 to 10 years depending on trap coverage and the threshold target (whether Mf or ATP). By contrast, this reaches as high as between 10 to 20 years in the case of the heterogeneous fly biting scenario as shown in (b). Note that the y-axis is shown on a log scale. The study site is Masaloa. The killing efficacy of traps is 70% to 80%.

The results further show that increasing trap coverage from 40-50% to 60-70% will have only a moderate impact on the number of years saved under the homogeneous biting scenario, particularly in the case when the 1% Mf threshold is used (Figure 8(a)). By contrast, increasing EWT coverage will add substantially to the number of years saved for both thresholds when the fly population is heterogeneously distributed in a setting (Figure 8(b)). These results are for Masaloa but similar patterns are also observed for all four study villages (Tables S10-S14).

### Impact of EWT and MDA intervention scenarios on elimination and recrudescence probabilities

Table 2 summarizes the estimated probabilities of achieving either of these outcomes firstly when both annual MDA and switch MDAs alone are stopped once the 1% Mf threshold is crossed, and no other interventions are introduced. The results highlight that stopping these MDAs after the 1% Mf threshold is crossed will result in zero probability of transmission elimination during the immediate 5-year post MDA stoppage period that corresponds to WHO’s recommended PTS period. Following this period, however, elimination probabilities will increase gradually, such that by year 15 post MDA stoppage, these will reach moderately high levels ranging from 15-47% (Table 2). Intriguingly, a similar pattern of increasing recrudescence of infection is also observed as the simulation period extends from 5 years to 15 years post stoppage of MDA. The results indicate that this negative outcome (increasing recrudescence of transmission over time post stoppage of MDA) is a direct function of the fact that the fraction of transient model curves (as noted by the figures following the ∼ sign in the table) that arise immediately following the crossing of the 1% Mf threshold in each study site will decay slowly over time, with larger fractions of these models either declining gradually to zero or exhibiting positive resurgences by year 15 post MDA stoppage (Table 2).

**Table 2.** Transmission elimination and recrudescence probabilities estimated for each study site 5 and 15 years after stoppage of annual and switch MDA regimes. The transient behavior as described in the methods section is indicated by the ∼ sign.

| Study Villages<br>(baseline Mf<br>prevalence (%)) | Annual MDA upto 1% WHO threshold |  |  |  |  |
| --- | --- | --- | --- | --- | --- |
|  |  | Elimination probability (%) |  | Recrudescence probability (%) |  |
|  | Time to cross<br>threshold<br>(year) | After 5 years | After 15 years | After 5 years | After 15 years |
| PalaurePacunaci<br>(100) | 25 | 0 (~68) | 42 (~14) | 32 | 44 |
| Masaloa (76) | 20 | 0 (~72) | 47 (~24) | 18 | 29 |
| Nyimanji (58) | 18 | 0 (~80) | 36 (~26) | 20 | 38 |
| OlimbuniAroga<br>(24) | 15 | 0 (~89) | 41 (~31) | 11 | 28 |
|  | Annual MDA upto 10 years then Biannual MDA upto 1% WHO threshold |  |  |  |  |
|  |  | Elimination probability (%) |  | Recrudescence probability (%) |  |
|  |  | After 5 years | After 15 years | After 5 years | After 15 years |
| Palaure Pacunaci | 18 | 0 (~73) | 25 (~27) | 27 | 47 |
| Masaloa | 15 | 0 (~88) | 29 (~39) | 12 | 32 |
| Nyimanji | 14 | 0 (~85) | 19 (~42) | 15 | 39 |
| Olimbuni Aroga | 11 | 0 (~93) | 15 (~57) | 7 | 28 |

However, while these patterns are found to occur irrespective of whether annual MDA or switch MDA was simulated, an interesting finding was that annual MDA performs better than the more intensive switch MDA strategy for inducing longer term (year 15) transmission elimination whilst correspondingly reducing recrudescence of infection in each site. This appears to be mainly due to the fact that annual MDA generates lesser transient dynamics in the model outcomes compared to switch MDA (Table 2). Finally, there is some indication that the long-term probability of resurgence or recrudescence of infection may be related positively with baseline Mf prevalence, increasing as baseline prevalences increase.

The corresponding effects of including EWT post-stoppage of MDA when deployed monthly either during the peak biting season or throughout the year for a moderate level of mean trap fly killing efficacy (75%) and coverage (65%) configuration on transmission elimination/recrudescence are shown for each of the four villages in Tables 3-4 (see Tables S15, S16 for results for high efficacy and coverage EWT). The predictions indicate that in contrast to when EWT is not introduced (Table 2), the deployment of these traps following the stoppage of MDAs will dramatically increase long-term onchocerciasis transmission elimination whilst significantly suppressing the probability of recrudescence over both the short as well as longer-terms. Thus, the inclusion of the traps will generate very high elimination probabilities (up to as high as 97%) but only if they are deployed over the long-term (15 years in the present simulations), with the period immediately post stoppage of MDA marked by significant transient dynamics (with between 96% to 100% of curves exhibiting this feature during the first 5 years post MDA stoppage). Importantly, these results do not differ appreciably whether monthly EWT is deployed only during the peak biting season or annually (compare Table 3 versus 4), although slightly higher probabilities of transmission elimination result when traps are deployed monthly. Increasing the fly killing efficacy and coverage of EWT to levels as high as a mean of 95% for each of these control parameters will have the strongest impact in reducing the recrudescence probability to 0 at both 5- and 15-years post MDA, while increasing the long-term probability of transmission elimination to as high as 99% (Tables S15, S16). This latter finding implies that deploying just 7-8 EWT traps during the peak biting season in a village of size 1 *km*^2^ will be sufficient to suppress high probability recrudescence and achieve long-term high probability transmission elimination. A further notable feature of the results displayed in Tables 3 and 4 is that the deployment of EWT after the stoppage of MDA may suppress resurgence of infection uniformly across all the study sites, i.e. irrespective of baseline infection prevalences. These results are for the homogeneous fly biting case; except for increased timelines to cross the 1% Mf threshold, qualitatively the same elimination/recrudescence patterns were also observed for the heterogeneous biting case.

**Table 3.** Elimination and recrudescence probabilities estimated for each study site 5 and 15 years after stoppage of the switch MDA regime (10 years annual and then biannual MDA for the remaining time until the 1 % Mf threshold is crossed). Here EWT fly killing efficacy is 0.75 (0.7 − 0.8) while coverage is 0.65(0.6 − 0.7). EWT is deployed monthly during the peak biting season. Transient behavior of models as described in the methods section is indicated by the ∼ sign.

| Study Villages | Annual MDA upto the 1% WHO threshold followed by monthly EWT during the peak biting season |  |  |  |  |
| --- | --- | --- | --- | --- | --- |
|  |  | Elimination probability (%) |  | Recrudescence probability (%) |  |
|  | Time to cross threshold (year) | After 5 years | After 15 years | After 5 years | After 15 years |
| Palaure Pacunaci | 25 | 0 (~96) | 85 (~8) | 4 | 7 |
| Masaloa | 20 | 0 (~100) | 94 (~5) | 0 | 1 |
| Nyimanji | 18 | 0 (~100) | 92 (~7) | 0 | 1 |
| Olimbuni Aroga | 15 | 0 (~100) | 95 (~4) | 0 | 1 |
|  | Annual MDA for 10 years plus Biannual MDA upto the 1% WHO threshold followed by monthly EWT during the peak biting season |  |  |  |  |
|  |  | Elimination probability (%) |  | Recrudescence probability (%) |  |
|  |  | After 5 years | After 15 years | After 5 years | After 15 years |
| Palaure Pacunaci | 18 | 0 (~98) | 79 (~15) | 2 | 6 |
| Masaloa | 15 | 0 (~100) | 88 (~11) | 0 | 1 |
| Nyimanji | 14 | 0 (~100) | 84 (~15) | 0 | 1 |
| Olimbuni Aroga | 11 | 0 (~100) | 85 (~15) | 0 | 0 |

**Table 4.**
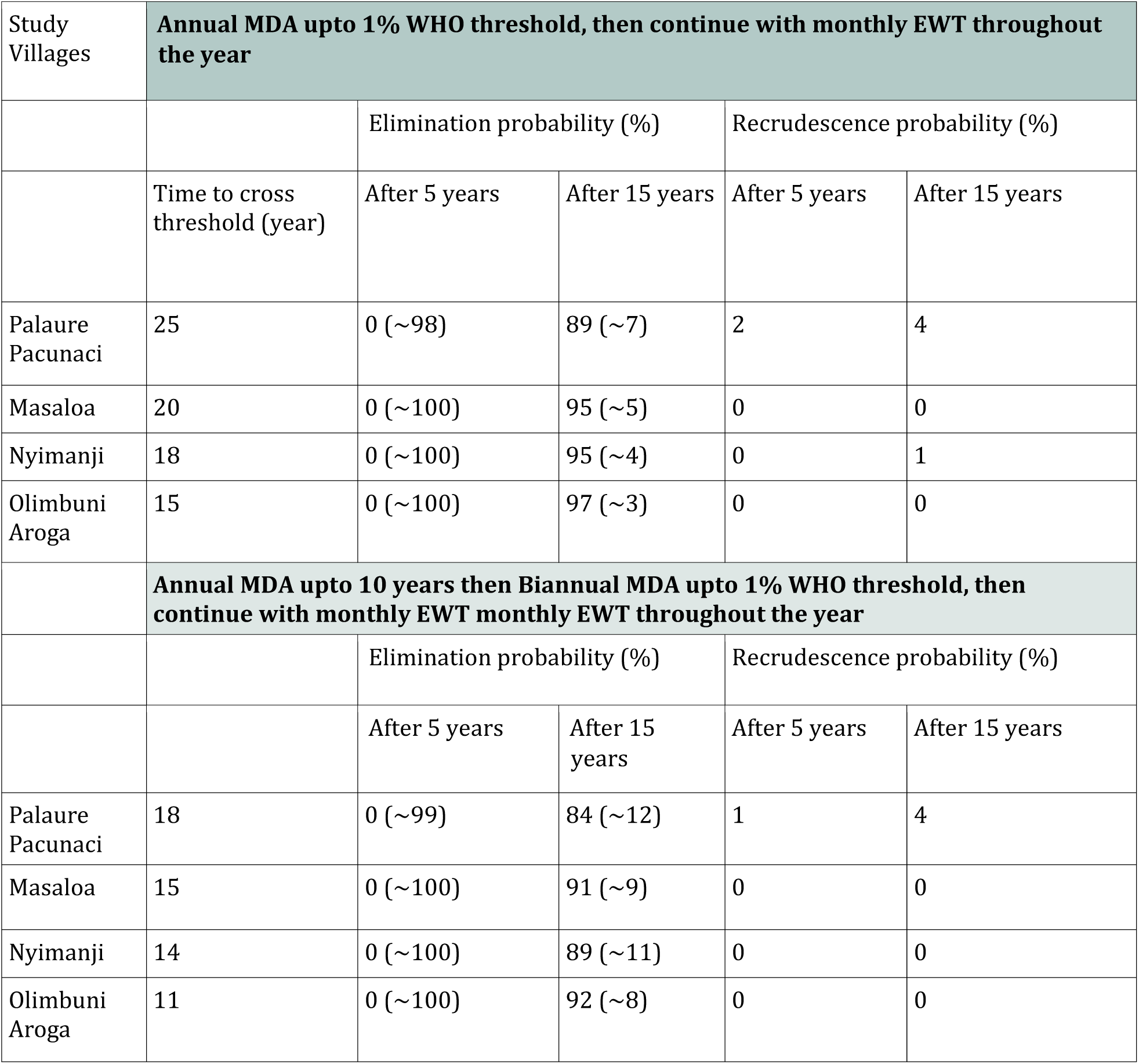
Elimination and recrudescence probabilities estimated for each study sites 5 and 15 years after stoppage of switch MDA regime to reach 1% Mf threshold (10 years annual and then biannual for the remaining time to reach 1 % Mf threshold). Here EWT killing efficacy is 0.75 (0.7 − 0.8) while coverage is 0.65(0.6 − 0.7). EWT is deployed monthly throughout the year. The transient behavior as described in the methods section is indicated by ∼ sign.

## Discussion

The Esperanza Window Trap (EWT) is a recent addition to community-directed VC approaches which empirical field studies have shown could serve as a means for bringing about significant reductions of *Onchocerca*-transmitting black flies (25, 26, 28, 30, 31). Further, it has been demonstrated that EWT can be deployed effectively by community members such that it could also constitute a measure for overcoming the problems of implementing the sustained long-term black fly control required to meet the endgame challenges of achieving onchocerciasis eradication (21, 34). However, currently no study exists where the combined effects of MDA and EWT have been investigated for reducing infection in both the vector and human populations to levels that can result in the achievement of community-wide transmission elimination of the parasite. EWT deployment at the village/site scale for achieving black fly control also requires understanding of the factors that will underlie the configuration of the trap in the field, including the density of traps and where to place them, needed to bring about optimal area-wide fly reduction in a given landscape (33). Here we have sought for the first time to shed light on this central issue connected to the significance of using EWT as a transmission elimination tool by adapting and coupling a population-level model of onchocerciasis transmission with models of various EWT trap networks deployable in the field. More specifically, we used the developed trap network-onchocerciasis transmission modeling system to address how operational parameters connected with the field deployment of EWT, such as efficacy of traps in trapping and killing *Simulium* flies, number of traps and coverage of a setting, inter-trap distance, capture radius, and heterogeneity in fly distribution in a community (33, 58), may affect the abundance of the Simulid fly population, and in combination with reductions in human infection brought about by MDA, can contribute to the community-wide elimination of onchocerciasis transmission.

Our first analysis in this work examined the feasibility of using EWT to eliminate parasite transmission from the black fly population in a community, focusing on the number of traps required to achieve this goal under various trap network parameters, transmission conditions, and areal settings (Figure 4, Table 1). Our simulations highlight that while the ATP threshold of 20 infective bites per person per year may indeed be effectively breached quickly (even within a year) through the monthly deployment of EWT traps when these are arranged in a rectangular array in a community (the results shown in Figure 4 and Table 1 reflecting the outcomes predicted for the meso-endemic village of Masaloa although similar results were obtained for the other three study sites), the numbers of traps required to actually achieve this goal will vary as a complex function of trap fly killing efficacy, trapping radius, inter-trap distance, fly biting heterogeneity and size of a setting. Thus, for example, if the trapping radius of a trap and inter-trap distance are large (such as the 200m and 490-564m investigated here), the number of traps given a homogeneous fly biting scenario and a fly killing efficacy of 70-80%, for achieving the ATP threshold by means of a moderately high trap coverage (60- 70%) and monthly trap placements for a year, can vary from as little as just 5 in a small size setting (1 *km*^2^) to as high as 487 in the case of a large size community (10 *km*^2^) (Table 1). These numbers will increase with coverage, eg. to 8 and up to 796 for this same scenario if the trap coverage is increased to levels as high as 90- 100%. A key finding, however, is that as spatial heterogeneity in fly biting increases, the number of traps required to achieve the same goal will decrease significantly compared to the homogeneous biting case, although even at the highest trap radius/inter-trap distance studied here up to 98 traps will still be required (at the 60-70% coverage level) to cross the ATP threshold at the most heterogeneous situation modelled (ie. 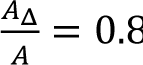) in the largest area examined here (10 *km*^2^) (Table 1). The results displayed in Table 1 also indicate that the number of traps needed to achieve the ATP threshold is highly sensitive to trap capture or fly attractant radius and may become infeasibly high if this radius, and hence the inter-trap distance, decreases to low levels.

Another important result in this regard which has major implications for the design of effective EWT trapping networks is that the trap coverage (and therefore number of traps) needed to cross the 20 ATP threshold via placement of traps every month for a year is found to depend strongly on the trap fly killing efficacy (Figure 4d). Thus, while an efficacy of 70% will require a trap coverage of just over 60% for a trap network consisting of traps with a capture radius of 200m, placed every 490m, and deployed in a setting of size 1 *km*^2^, much higher coverages will need to be achieved to meet the goal of achieving this target within a year if the fly killing efficacy drops below 70% (Figure 4d). Indeed, there is a critical fly killing efficacy (approximately 45%) below which the ATP threshold cannot be achieved within a year even at maximal coverage, indicating the strong sensitivity of the trap network to the fly killing efficacy of the trap.

These predictions highlight on the one hand the inadequacy of simply transferring evidence regarding fly killing effectiveness based on restricted field trials for evaluating the utility of using EWT for bringing about area- or community-wide interruption of transmission from the vector population. Indeed, our results show that such evaluations must be made only by considering the effects of a given EWT trapping configuration or network on parasite transmission dynamics, including the biting distribution of the vector, in a locality. These results also underscore that high trap coverages, and thus infeasibly high trap numbers, will be required to achieve vector-based elimination targets rapidly, which might limit the use of EWT traps if only much lower coverages can be attained from using fewer traps and longer inter-trap distances (see Table 1 for a comparison of the number of traps needed at 60-70% and 90-100% coverages for different trap capture radii). This, however, will lengthen the timelines required to attain the ATP elimination target used in the present simulations.

By contrast, the simulations of including EWT with MDA on the timelines required to reach the 20 ATP and 1% Mf thresholds investigated here using our MBR seasonality incorporated onchocerciasis transmission model demonstrate that both of these thresholds can be achieved faster by coupling MDA strategies with EWT compared to using MDAs alone (Figures 5-8 and Tables S5-S14). Overall, including EWT in the MDAs (for trap killing efficacy of 75%, capture radius of 200m and MDAs delivered at a coverage of 80% in a setting covering an area of 1 *km*^2^ and experiencing homogeneity in fly biting) can potentially save between 1-12 years across the 4 study sites for combined annual MDA/EWT and 1-17 years when EWT is combined with switch MDA (Table S5). A noteworthy feature of these results is also that the ATP threshold will be achieved much faster than the corresponding Mf threshold in each village setting (and therefore saving more intervention years if used as a target) when EWT is coupled with either of the MDA strategies irrespective of the biting fly distribution in a setting (Figure 8). As we have shown previously this outcome is primarily a function of the shorter life span of parasite larval stages and greater susceptibility of filarial vectors to VC measures compared to the markedly longer life expectancy and comparatively lesser susceptibility of the adult parasite populations residing in human hosts to drug treatments (34, 35). Note also, as we have highlighted previously (34), while this result indicates that targets based on infection indicators in the vector (ATP) may be significantly more sensitive for early detection of transmission interruption, MDA will still be important for reducing the intensity of the remaining worm infections in humans in order to achieve the permanent reduction of transmission especially in settings where in-migration of black flies is thought to be very likely.

A related significant finding is that while the frequency of using EWT (viz. a month before peak fly biting season, during the three months of the peak biting season, and monthly throughout the year) has little impact on the predicted timelines required to achieve the Mf threshold, using monthly deployment of EWT alongside the two MDAs modelled here by contrast can significantly reduce the years required to breach the ATP threshold compared with using MDA alone (Figures 5-8; Tables S5-S14). This outcome was observed irrespective of the fly biting variation, but an unanticipated finding is that times to reach both the 1% Mf and 20 ATP thresholds were found to increase for all interventions, including in the case of inclusion of EWT with MDA, as fly biting heterogeneity in a setting increases (Figure 7). This is because the spatial heterogeneity in vector biting as modelled here implies markedly higher than average densities of biting flies in certain sections of a community or village, which will accordingly translate to requiring longer times to breach the community-wide aggregated ATP and Mf elimination thresholds. On the other hand, high levels of vector control coverage can be achieved in the heterogeneous biting scenario by using lower number of traps as compared to the corresponding homogeneous setting. This is because placing the traps where flies are concentrated rather than everywhere saves resources in terms of trap numbers without compromising on coverage. However, a subtle but critical finding in this regard is that while increasing EWT coverage will only have moderate effects in the homogeneous biting case, achieving high coverages in those areas where most infective vector biting occurs will add substantially to the number of years saved for both thresholds (Figure 8(b). This finding will increase the feasibility of using EWT as a control measure for accelerating onchocerciasis elimination, but it will require baseline surveys of fly biting habitats within a setting to be performed before deploying an effective EWT configuration (number of traps, inter-trap distance) for a location of a specific size.

However, in general, the finding overall in this study of a nonlinear association between the number of traps required, fly biting distribution, coverage and areal size on timelines to reach the two thresholds studied for a given trap fly killing efficacy, capture radius and inter-trap distance means that: 1) trap numbers will not scale linearly with changes in areal size, ie. the number of traps used in a site will not translate simply to twice the number required for another site with double the area, and 2) the number of traps required will vary between sites with different areas even when they exhibit the same fly distribution pattern. This outcome again highlights the importance of considering the interaction between a given trap configuration and parasite population dynamics in settings varying in spatial fly biting patterns when evaluating the effectiveness of using EWT with MDA for progressing the meeting of set elimination targets.

Our third major result from this study pertains to simulations of the impact of adding EWT to MDA regimens for sustaining long-term transmission elimination once the community-wide 1% Mf threshold is crossed and MDA is stopped fully. This is becoming a critical topic of study given findings from both field and modeling studies which indicate that achieving these or similar thresholds may not lead to interruption of transmission in every setting (9, 60). More recent modeling studies also suggest that this outcome may intriguingly reflect the emergence of transient dynamical behavior near elimination thresholds, which could result in long-term low-level persistence of filarial parasitic systems (59). To imitate the current MDA interventions and these endgame questions that have emerged regarding ensuring that long-term elimination of transmission is sustained, we introduced EWT only after the 1% Mf threshold is crossed and MDA is stopped and again compared annual versus switch MDA strategies in this exercise. The results shown in Tables 2-4 and in Tables S15 and S16 highlight firstly the profound impact that introduction of EWT into either MDAs can have in enhancing the achievement of sustainable transmission elimination especially when EWT is implemented long-term compared to when no further interventions, including EWT, is introduced once MDAs are stopped. Thus, while long-term application (at least for 15 years) of EWT at even at moderate efficacy (75%)/coverage (65%) levels can result in in very high elimination probabilities (up to 94%) in the study villages when deployed monthly during the peak biting season (Table 3) and up to as high as 97% (much higher if higher efficacy and coverage levels are used (Tables S15, S16)) when used on a monthly basis (Table 4), the corresponding probabilities of transmission elimination achieved in the absence of EWT will only increase to 15-47% by year 15 post MDA stoppage with significant resurgences of infection predicted at both 5 and 15 years post MDA depending on the MDA regimen and village (Table 2). A further significant finding of import is that zero elimination is predicted for all the strategies with and without inclusion of EWT modelled here at year 5 post-MDA (Tables 2-4), largely because of the induction of transient transmission dynamics (majority of model curves exhibiting fluctuating low-level declines and increases in infection (59)) immediately following the crossing of the 1% Mf threshold.

These results imply firstly that while on the one hand deploying EWT even at moderate efficacy/coverage levels post MDA can increase onchocerciasis elimination probabilities whilst suppressing recrudescence of infection once the 1% Mf threshold is crossed compared to the situation when EWT is not implemented, this impact will only become apparent when EWT is implemented long-term after the crossing of this threshold. The finding of transient behavior immediately following the crossing of the 1% Mf threshold further implies that while infection levels may remain at low levels - even below the 1% Mf value - during the short term (for at least 5 years) after MDA is stopped, there will still be a possibility of significant resurgence of infection over the longer-term in the absence of VC measures, such as EWT. This outcome, ie a delayed resurgence of infection once the transient period passes, has major implications for post elimination surveillance. It suggests that such surveillance will need to be extended beyond the currently proposed PTS/PES periods (43) to facilitate a more reliable assessment of transmission stoppage in a setting especially when no VC measures are deployed during the post-MDA period (59). A final point of interest regarding the results arising from the deployment of EWT after the stoppage of MDA is that it will also overcome the impact of baseline endemicity levels by suppressing the resurgence of infection uniformly across sites differing in these initial transmission conditions (Tables 3, 4). As remarked previously (36, 46, 61), this outcome indicates how including VC measures during the endgame phase of an elimination program can have the additional important effect of overcoming the inherent between-site heterogeneity in parasite transmission dynamics, affording thereby a more reliable strategy for bringing about filarial transmission elimination across disparate localities.

On a more epistemological level, our results have provided additional elaborations of the role that mathematical models which can integrate elements of an intervention with parasite transmission dynamics can play in generating genuinely new knowledge regarding how a specific intervention would serve to meet a set population-wide disease management goal (45, 62). In particular, here, we have demonstrated how primary information or hypothesis about the efficacy of EWT traps in reducing the black fly population obtained through focused field observations can be extended through our coupled EWT-onchocerciasis transmission model into producing higher-order insights regarding the use of EWT traps in a community for meeting set elimination thresholds, and following this for bringing about long-term sustainable transmission elimination. The new information gleaned from these simulations regarding the specifics of the trap network or configuration (number of traps, impacts of fly killing efficacy, trap capture radius, inter-trap distance and placement of traps over space and time taking into account vector biting distribution and seasonality) represent critical variables for meeting these goals effectively, which are often not fully explored or are challenging to examine using empirical studies alone. This ability of mathematical models, especially when used in conjunction with methods (such as the BM methodology used here) to assimilate information regarding local transmission conditions, to extend the evidence from one set of observations, ie. from field studies regarding the effectiveness of the EWT for reducing fly abundance at a particular spatial and temporal scale, for the generation of novel higher-order out-of-sample predictions about other variables and processes related to the overall operation of a dynamical system via the structures and parameters of a transmission model indicates on the one hand the limitations of making inferences based on restricted empirical data alone. Conversely, the results underscore the value of coupling structurally appropriate parasite transmission models with such data for allowing a more comprehensive and reliable assessment of the potential of an intervention, including guiding its optimal design, for facilitating community-wide parasite control across disparate endemic settings.

Our coupling of EWT trap network configurations with the seasonal MBR-based onchocerciasis transmission model has also presented new modeling constructs for how to incorporate the effects of variable spatio- temporal processes into a population-level vector borne macroparasite transmission model. In particular, we show how rainfall-driven seasonality in black fly biting abundance and how spatial heterogeneity in such abundances that may occur in landscapes of varying areal sizes can be structurally integrated with a population-level onchocerciasis model to gain more realistic insights of the impacts of control efforts to achieve the elimination of the transmission of *O. volvulus* in the field. The formulations by which we incorporate these heterogeneities into our base onchocerciasis transmission model as described in this paper (see specifically the text around Equation 8) thus constitute major innovations in enhancing the capabilities of data-driven population-level deterministic models for advancing investigations of the complex transmission and control dynamics of macroparasitic diseases in the real-world. We indicate that similar extensions could also be conceptualized and implemented into population-level parasite transmission models for other macroparasitic diseases, which would allow leveraging of their higher tractability and simplicity (as compared to stochastic or individual-based models) whilst improving their capability to handle the complex spatio-temporal heterogeneities, such as those addressed in this work, that are very likely to underlie the real-world disease transmission and extinction dynamics in the field.

A limitation to our study is that the exact form of the potential fly biting distribution in communities is not addressed explicitly. The general principles regarding trap placement and numbers given trap fly killing efficacy and coverage discussed in this paper can be expanded to include more realistic spatial distribution functions for the biting black fly population, although the analysis will be mathematically involved (33). The most common approach to trap deployment, as modelled here, is to place traps on a regular grid of cells across a spatial domain, but if sites most likely to result in catches of established or invading black flies are known, then it might be possible to set the traps in these optimal fly habitats. Research studies will be required to determine such optimal habitats for trap placement to take advantage of the lower trap numbers that would be required in such circumstances. Furthermore, given that the number of traps required to form an effective attractant-based trap network is strongly sensitive to trap insect-killing efficacy and fly capture radius, future empirical work to quantify these parameters for different lures, including how these will change as lures age, is needed. The movement of flies over a landscape is another area where the present modeling system can be made more realistic. If better data becomes available on black fly dispersal behavior, then it might be possible to include a movement model for the fly population in our EWT simulation system (63, 64), although if the dispersal behavior is also affected by host physiology and mobility as well as by habitat patchiness in a setting then it might call for the extension of the present models into more spatially-explicit simulation systems, such as the agent-based vector-borne disease modeling frameworks being investigated for onchocerciasis by other authors (39). Finally, we have used elimination targets initially proposed by WHO (WHO, 2016) in all the simulations carried out in this work. Our previous modeling studies have indicated that such breakpoints are not global, can vary by endemicity, and may be lower than the thresholds used here (34, 46, 50), although note that the present threshold values may apply in the presence of black fly control (34, 46). While this indicates a need for reevaluating and confirming the targets required for determining whether transmission has been interrupted, the key conclusions of our study, viz. that adding EWT VC to MDA will significantly reduce timelines to elimination targets, and following this, will enhance the achievement of sustained long-term transmission elimination compared to MDA alone in communities will not change although the actual durations over which both goals will be met would (34, 46).

## Data Availability

All data produced in the present work are contained in the manuscript

https://github.com/EdwinMichaelLab/OnchoEWT

## Acknowledgment

E.M. and T.R.U. thank the National Institute of Allergy and Infectious Diseases (https://www.niaid.nih.gov/) for financial support of this study (Grant RO1AI123245). The funders had no role in study design, analysis, decision to publish, or preparation of the manuscript.

## Author Contributions

Conceptualization: E.M. and S.B. Formal Analysis: S.B., S.S., W.Z.; M.E.S., E.M. Provided Data: T.R.U. Code Organization: K.N. Writing- original draft: S.B. and E.M. Writing – review and editing: All authors.

## Supporting Information Captions

**S1 Fig.**
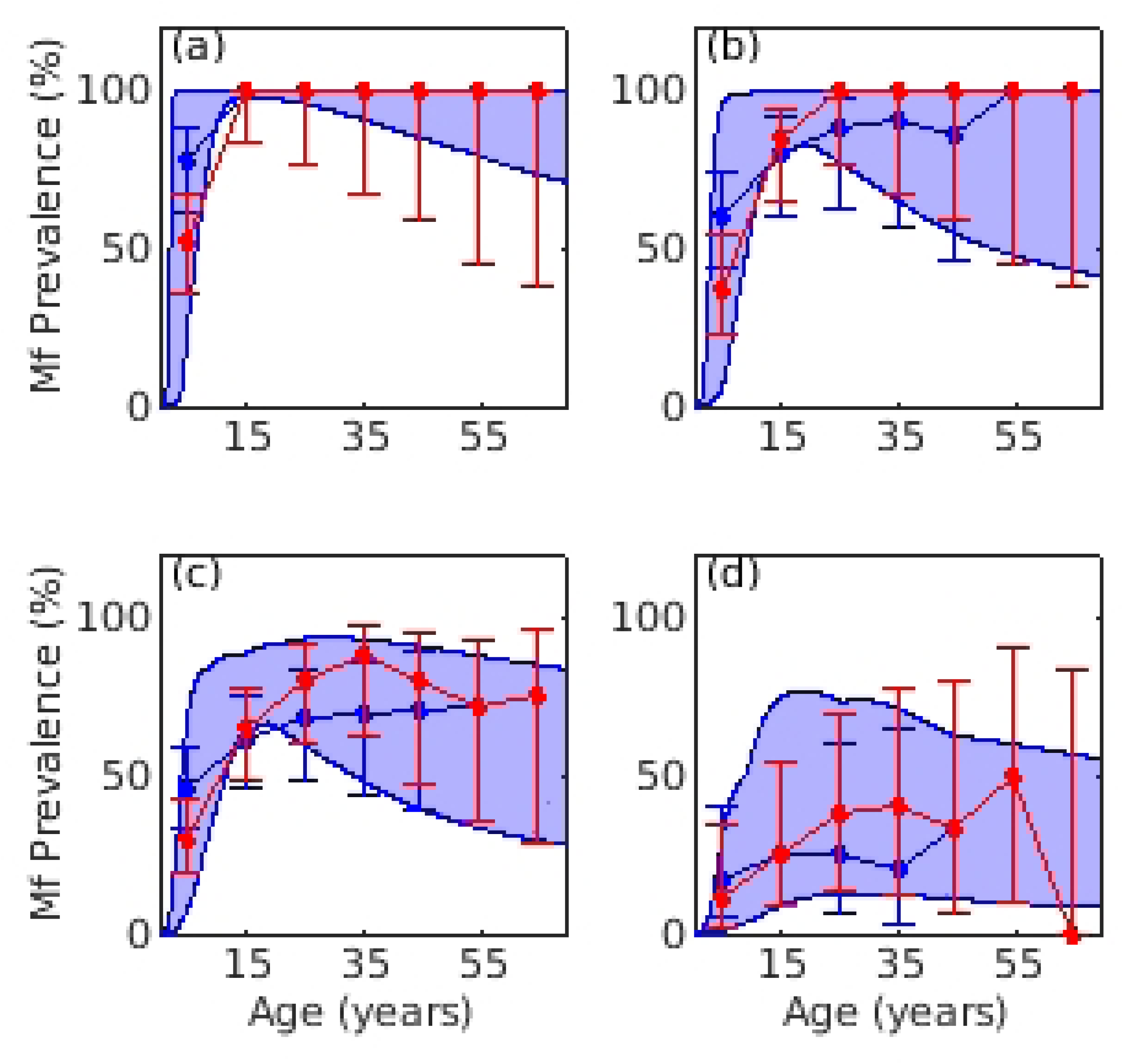
Model fits to baseline age-prevalence in study sites.

S1 Table. Parameters of the onchocerciasis transmission model.

S2 Table. Details of age and population dependent functions used in the transmission model.

S3 Table. Baseline Mf prevalences in the four study sites.

S4 Table. Priors of parameters related to trap efficacy, decay rate, drug efficacy, and for terms describing seasonal MBR.

S5 – S9 Tables. Years predicted for reaching the WHO set thresholds in the four study sites under annual and switch MDA with and without EWT in the homogeneous fly biting condition.

S10 – S14 Tables. Results on predicted timelines to reach the WHO set thresholds in the four study sites under annual and switch MDA with and without EWT in the heterogeneous fly biting condition.

S15 – S16 Tables. Estimated transmission elimination and recrudescence probabilities arising from the deployment of EWT at high coverage and efficacy values post stoppage of MDA during months of peak biting season and monthly throughout the year.

## Code availability

All the Matlab codes used in this work are available in Github: https://github.com/EdwinMichaelLab/OnchoEWT

